# Effectiveness of shared medical appointments delivered in primary care for improving health outcomes in patients with long-term conditions: a systematic review of randomised controlled trials

**DOI:** 10.1101/2022.03.24.22272866

**Authors:** Mei Yee Tang, Fiona Graham, Amy O’Donnell, Fiona Beyer, Catherine Richmond, Falko Sniehotta, Eileen Kaner

## Abstract

**Objectives:** Shared medical appointments (SMAs) have the potential to address interlinked challenges of limited capacity in primary healthcare and rising prevalence of patients with multiple long-term conditions (LTCs). This review aimed to examine the effectiveness of SMAs compared to one-to-one appointments in primary care at improving health outcomes and reducing demand on healthcare services.

**Methods:** We searched for randomised controlled trials (RCTs) of SMAs involving patients with LTCs in primary care across six databases from 2013-2020 and added eligible papers identified from previous relevant reviews. Data were extracted and outcomes narratively synthesised, meta-analysis was undertaken where possible.

**Results:** Twenty-three unique trials were included. SMA models varied in terms of components, mode of delivery and target population. Most trials recruited patients with a single LTC, most commonly diabetes (*n*=12), although eight trials selected patients with multiple LTCs. There was substantial heterogeneity in outcome measures which we categorised into health outcomes (biomedical indicators, psychological and well-being measures), healthcare utilisation, and cost and resource use. Meta-analysis showed that participants in SMA groups had lower diastolic blood pressure than those in usual care (*d*=-0.123, *95%CI* = - 0.22, -0.03, *n=*8). No statistically significant differences were found across other outcomes. Compared with usual care, SMAs had no significant effect on healthcare service use.

**Conclusions:** SMAs were at least as effective as usual care in terms of health outcomes and did not lead to increased healthcare service use in the short-term. To strengthen the evidence base, future studies should target standardised behavioural and health outcomes and clearly report SMA components so key behavioural ingredients can be identified. Similarly, transparent approaches to measuring costs would improve comparability between studies. To better understand SMA’s potential to reduce demand on healthcare services, further investigation is needed as to how SMAs can be best incorporated in the patient care pathway.

**PROSPERO protocol registration:** *CRD42020173084*

299/300

**STRENGTHS AND LIMITATIONS:**

- Focus on randomised controlled trials, highest quality evidence of the effectiveness of SMAs in primary care for long term conditions
- Robust search strategy, based on previous high-quality review; refined by information specialists to focus on primary care
- Rapidly evolving area of practice and publications and the most recent evidence may be missing.
- Small number of studies reported resource use and costs limiting conclusions regarding efficacy of SMAs in primary care.

**FUNDING:** This paper is independent research commissioned and funded by the NIHR PRU in Behavioural Science (Award: PR-PRU-1217-20501). The views expressed in this publication are those of the authors and not necessarily those of the NHS, the National Institute for Health Research, the Department of Health and Social Care or its arm’s length bodies, and other Government Departments. The funders had no role in the design of the study, collection, analysis or interpretation of data or in the writing of the manuscript.

**COMPETING INTERESTS:** None to declare.

**CHECKLIST:** See supplementary material for PRISMA checklist.

## INTRODUCTION

Shared Medical Appointments (SMAs), also known as group consultations, are a model of care with the potential to address the interlinked challenges of limited capacity in primary care and rising prevalence of patients with multiple long-term conditions (LTCs)[1,2]. SMAs are longer appointments (typically 60-120 minutes) whereby patients with the same LTCs meet with their physician together[3]. SMAs are typically co-led and/or facilitated by healthcare professionals, such as nurses, pharmacists, psychologists, and physiotherapists. The group typically consists of between 6-15 patients and may include family members and caregivers[4,5]. There are various models of SMA but generally they retain some features of a standard one-to-one appointment such as physical examinations and personalised review of medical charts[2]. In addition, SMAs provide participants an opportunity to ask questions of clinicians and other patient and receive formal education and counselling during the group session. SMAs have been used to deliver care for a range of health conditions including diabetes, hypertension, and chronic pain; though there is potential for wider application, including multimorbidity[6].

A recent synthesis of qualitative literature found that most patients and primary care practitioners regarded SMAs positively[6]. Key benefits included improved patient self-confidence and motivation for self-management; whilst practitioners felt that SMAs had the potential to provide a more efficient and effective way of delivering care[7].Previous reviews of effectiveness were inconclusive but evidence, largely from the United States (US) and Australia, reported a promising effect of SMAs for some biomedical measures. For example, improvements in glycated haemoglobin A1C (HbA1C) and systolic blood pressure (SBP) were greater in patients attending SMAs compared to usual care for diabetes[8,9]. However, previous reviews include studies that evaluate the use of SMAs in secondary care settings as follow-up appointments to specialist treatment [4]. A mixed-methods review of SMAs in primary care settings for non-specialist treatment concluded that SMAs can yield improvements in patient satisfaction and some biophysical markers of disease [5]. However, this review conducted in 2015 included studies of SMAs for non-LTCs. It remains unclear whether SMAs are effective in supporting improved ongoing management of LTCs in primary care.

This review examined the effects of SMAs delivered in primary care on health outcomes and healthcare service use in patients with LTCs. We sought to answer two overarching research questions:

1. **Are SMAs effective in improving health outcomes for patients with one or more long-term conditions?**
2. **Do SMAs reduce healthcare service use by patients with one or more long-term conditions?**

## METHOD

This systematic review follows Cochrane Handbook Guidance[10] and the Preferred Reporting Items for Systematic Reviews and Meta-Analyses (PRISMA) guidelines[11].

### Protocol and registration

This study was registered on PROSPERO (<u>CRD42020173084</u>). Regarding protocol changes, we proposed coding of Behaviour Change Techniques Taxonomy V1 (BCTTv1)[12] used in SMAs and associated with changes in outcomes. However, most included studies did not report the required detail and so instead we narratively described this information.

### Inclusion/exclusion criteria

Studies were included if they met the criteria outlined in Table 1. The focus of this review was on models of SMAs that include one-to-one time for every patient in attendance as per the description of group consultations reported previously [1,2].

**Table 1:**
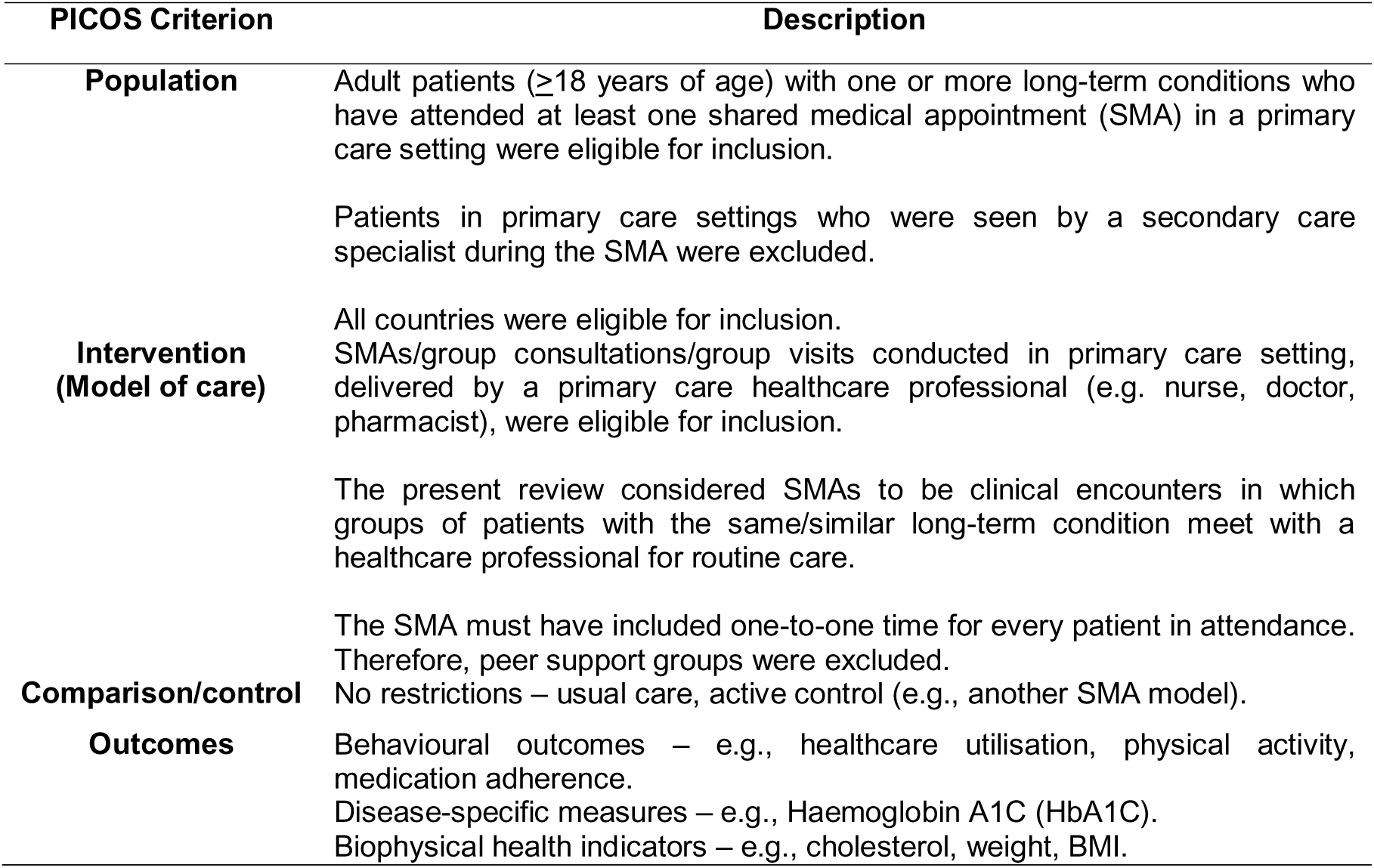
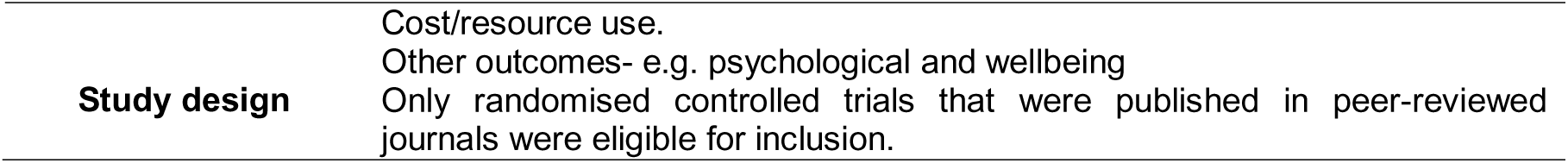
Inclusion and exclusion criteria.

### Search strategy

A comprehensive search strategy was developed, based on the approach described in Booth et al.[13], to search for trials published after their search, namely the period 2013-2020. Key changes included the removal of the terms “group outpatient”, “GMV” or “GMA”, “group processes” and “Group care” to improve the sensitivity and specificity of the search (see S1). The search strategy was first used to search MEDLINE (via OVID) and then translated for the following databases: EMBASE (via OVID), Science citation index (via Web of Knowledge) Social Science Citation Index (via Web of Knowledge), Cumulative Index to Nursing and Allied Health Literature (via EBSCOhost), Cochrane Central Register of Controlled Trials (Wiley), DARE, NHS EED, and HTA (Centre for Reviews and Dissemination). Any relevant pre-2013 trials identified by forward and backward citation searches of the included trials, including those in relevant systematic reviews[4,6,14,15], were also included in the review.

### Screening

Screening and data extraction was facilitated using the systematic review management tool, Covidence[16]. Two reviewers independently screened all titles and abstracts against the inclusion criteria, and a third reviewer adjudicated any disagreements. This process was also applied to the screening of full-text papers.

### Data extraction

Information relating to the study design, population, and intervention were extracted based on a framework on form of delivery[17] (e.g. experience/training of the providers and facilitators), outcomes, and results were extracted from all relevant papers using a data extraction form (see S2). All information was double-extracted by two researchers, with disagreements resolved through discussion or third-party moderation.

Where data were reported for several time-points, the data-point closest to the end of the SMA intervention was used to calculate the effect size as this would be when the largest effects attributable to the SMA is expected. If available, intention-to-treat data were used to calculate effect sizes.

### Quality assessment

Two researchers independently assessed the quality of all included studies using the Cochrane Risk of Bias Tool[18]. Percentages of judgements (high, low, or unclear) for each domain was calculated across the studies.

### Data analysis/synthesis

We mapped all reported outcomes into the following categories agreed by the wider research team to best reflect SMA effectiveness and efficiency: health outcomes (biomedical indicators, psychological and well-being measures), healthcare utilisation and cost and resource use.

Meta-analyses were performed in StataIC 15. Given the heterogeneity between studies, a random-effects model was used[19]. Meta-analyses were conducted where there were at least two studies reporting a specific outcome[20]. Outcome effect sizes were calculated as Cohen’s *d* (standardised mean difference). Heterogeneity was assessed using Higgins I-Square (*I*^2^), whereby 50-90% was considered as representing substantial heterogeneity[21]. Authors were contacted for additional information if data needed to calculate effect sizes were not sufficiently reported in the published paper(s). Where this information could not be obtained from authors, *p*-values and confidence intervals were used to calculate effect sizes[10]. Only studies in which the comparator was usual care were pooled into the meta-analysis.

Using meta-regression, sensitivity analyses were conducted to explore whether results differed according to sources of bias identified from the risk of bias assessment.

Studies that were too heterogeneous to perform meta-analysis, and where the comparator was not usual care, were synthesised narratively. Extracted data was tabulated by outcome measure to enable comparisons and relationships across studies to be more easily examined [22]. For each outcome measure, evidence of an effect was determined by the *p*-values reported in the papers. To assess the certainty of the evidence, number of study participants, confidence intervals and the consistency of effects across studies, the risk of bias of the studies, how directly the included studies address the planned question (directness) were taken into consideration.

### Patient and Public Involvement

The PRU BehSci Public Patient Involvement group provided their patient perspective about outcome measures of interests.

## RESULTS

**Figure 1:** PRISMA diagram of study selection process

### Characteristics of the included studies

Twenty-three unique trials (reported in thirty-four papers) were identified, for PRISMA details see Figure 1 and Table 2. See S3 for list of included papers.

**Table 2:**
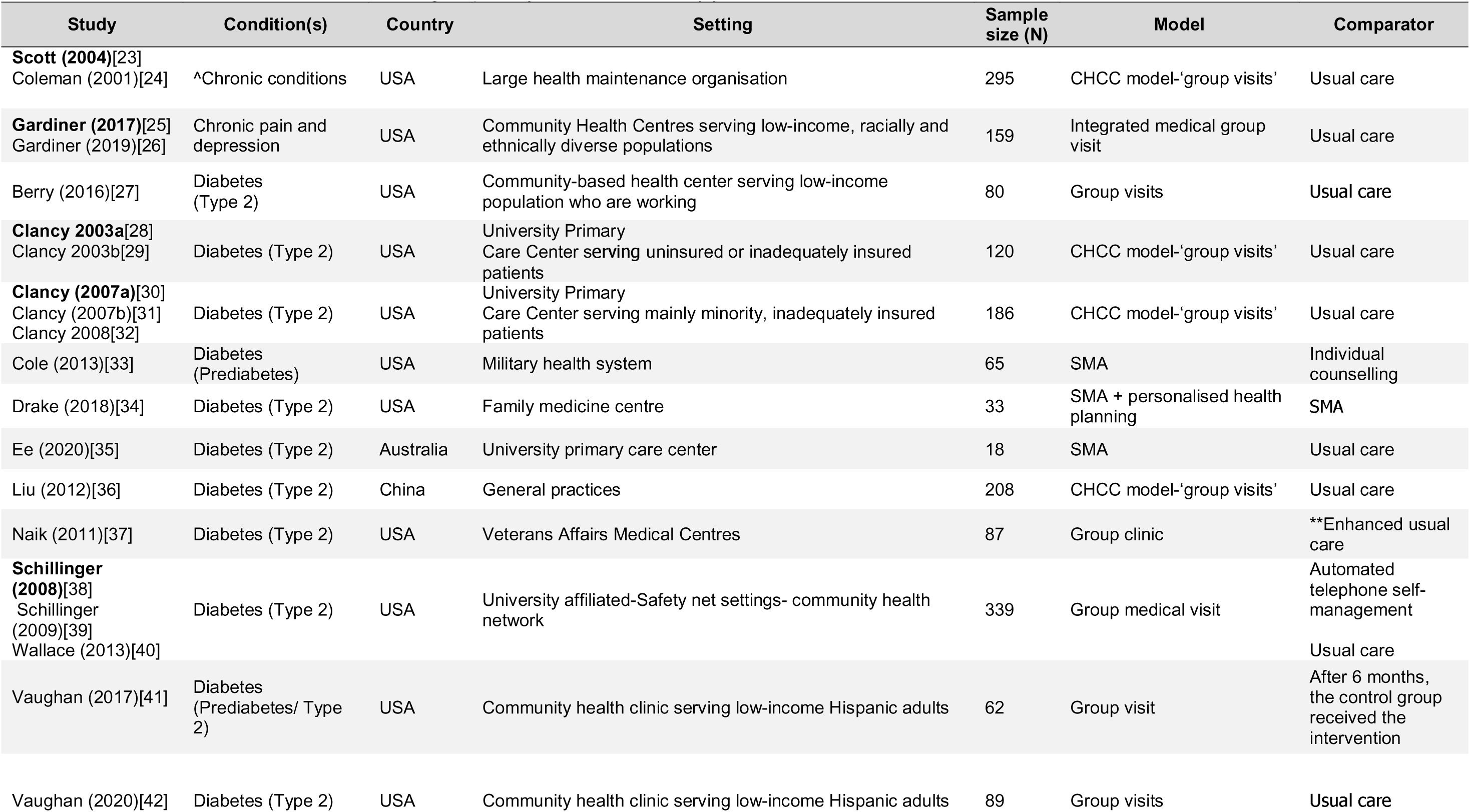
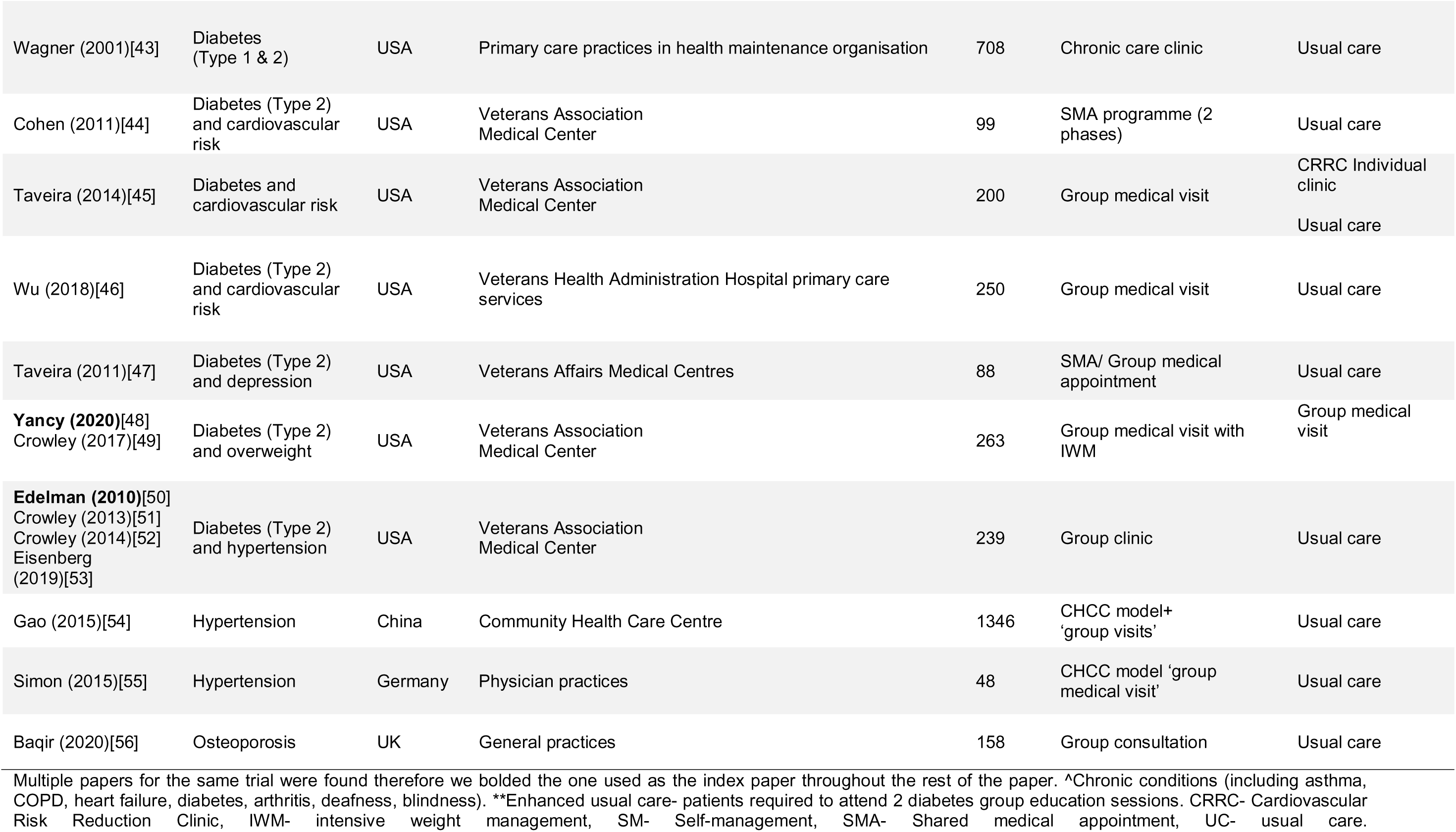
Characteristics of included studies grouped by health condition(s)

Fifteen trials (70%) were for a single LTC, of these: 12 were for diabetes[27,29,30,33–38,41–43], two for hypertension[54,55]; and one was for osteoporosis[56]. Eight trials considered multiple LTCs: three were diabetes and hypertension/cardiovascular risk[44–46,50]; one was diabetes and depression[47], one was for overweight patients with diabetes [48]; one was chronic pain and depression[26]; and one included multiple LTCs including: arthritis, hypertension, cancer, deafness and diabetes[23]. Overall, 20/23 (87%) of trials focused on patients with diabetes.

Eighteen trials (83%) were conducted in the US[23,26,27,29,30,33,34,37,38,41–48,50], two in China[36,54] and one each in Australia[35], Germany[55] and the UK[56]. Eleven trials were measured the effectiveness, impact or efficacy of SMAs compared to usual care[23,26,27,30,33,36,37,42,43,46,50], Eight trials examined feasibility parameters[28,34,35,38,41,42,47,54], and two trials were non-inferiority/superiority trials[48,49,56].

In eight trials (35%) participants were veterans or military personnel[33,37,44–48,50]. Participants were from low-income communities in four trials[26,27,41,42], and uninsured communities in three US trials[29,30,38]. Two trials were tailored for non-English speaking participants, where written materials were available in Spanish[38,42]. The majority of participants were over 50 years old, the mean age of participants ranged between 50.5[25,26] to 74 years old[56]. Two trials were specifically for older patients over 55 or 60 years respectively[23,55], and two trials excluded patients over 75[48] and 80 years[36]. Twelve trials (52%) had a majority of female participants[23,27,28,36,41,42,54,56]. Ten trials had majority of male participants[33–35,37,44–48,50]. One trial did not report the gender balance of participants[55]. Five studies had a majority White population[26,33,43,46,47], six trials had a majority Black population[27,29,30,34,48,50], two trials had a majority Hispanic population[41,42], two trials had a majority Asian population[35,36], and one trial had a majority White-Latino population [38]. Five studies did not report the ethnicity of participants[23,44,54–56].

Most trials (*n*=17, 78%) had a two-arm design that compared SMAs with usual care, typically a one-to-one (1:1) appointment with primary care physician[23,26,27,29,30,35,36,41–47,50,54,56]. In three other two-arm trials, the comparator was a 1:1 appointment plus two diabetes group education sessions[37], an SMA without an integrated weight management programme[48] or an SMA with a personal health planning component (PHP)[34]. Two trials had a delayed six-month waitlist control design[41,42]. Two trials had a three-arm design: one examined the effectiveness of a cardiovascular risk reduction clinic compared with group medical visits and usual care[45]; and a second compared an automated telephone self-management service with SMAs and usual care[38]. Where descriptions of usual care were available, usual care was delivered by a primary care provider, typically a physician/GP or nurse practitioner [57–62]and, in some cases, a pharmacist [62,63]or dietician [64]. A review of medication and chronic disease monitoring (e.g., measures of blood pressure and HbA1c) commonly took place in these sessions [58,59,63,65,66]. In some of the SMAs for diabetes, the usual care sessions included some form of individualised diabetes self-management education[58,62,64] or referrals were made available to see a diabetes educator dietician [59,67].

### SMA components and mode of delivery

There was much variation in the SMAs models reported by studies (see S9 for detailed description). Key features of SMA models were: facilitated group discussion or group question and answer session (15 trials)[19,22,25,26,29,30,32–34,37–39,40,43,46,50], ‘group education’ (14 trials)[19,23,25,31–33,37,38,42,43,44,46,50], and the opportunity to socialise (11 trials)[19,22,25,26,29,32,34,37,38,46,50].

SMAs were delivered face-to-face in all trials, although three SMA models included digital technologies, namely website and telephone support[26] and phone calls or text message support and/or reminders[41,42]. In three trials[37,45,56] SMAs were delivered by a single healthcare professional though mostly they were delivered by multidisciplinary teams. Professionals most commonly involved were family physicians[23,27,28,34–38,43,48,50,54,55], nurse practitioners[27,30–33,36,48,49] or nurses[23,24,43,44,46,50–54]. It was not always possible to tell what role each member of staff had in the delivery of the SMA. Provider characteristics other than profession or role were rarely reported, though two trials involving a majority Hispanic/Latino participants reported that the community health worker and or physician were bilingual[38,42]. Four trials reported that the same interventionists attended all visits for a particular group[44,48,50,55]. One trial of diabetes SMAs reported that group assignments were maintained for all SMAs to facilitate peer interactions and relationships within groups[37]. The consistency in group composition in terms of patient and interventionists attending each session was not reported by most studies.

Trial outcome measures varied across trials though most included biomedical indicators, psychological and well-being measures, healthcare service use and cost and resource use. Full details of all outcome measures reported by the studies are presented in S4.

### Risk of bias

Risk of bias item across the studies was generally low across the items, except for ‘Blinding of participants and personnel’ (83% of trials high, 17% unclear) (See S5).

#### Sensitivity analyses

There were no differences for any of the outcomes according to the risk of bias assessment criteria relating to random sequence generation, allocation concealment, blinding, incomplete outcome data, and selected reporting (see S5).

### Effectiveness of SMAs

#### Biomedical indicators

##### Glycated haemoglobin A1C (HbA1c) (%)

Of the 14 trials measuring HbA1c (%) which compared SMA to usual care, nine trials[27,30,35,38,42,45–47] were included in a meta-analysis. No statistically significant difference between SMAs and usual care was found for HbA1c (%) at follow-up (*d*=-0.098, *95%CI* = -0.34, 0.14, *n=*9) (*p*=0.420) (see Figure 2a). Substantial heterogeneity was observed (*I*^2^ = 70.9%). Of four other SMA trials reporting HbA1c (%) but not in the meta-analysis[29,43,44,50], only one reported significant between group differences, whereby the SMA group had significantly higher odds of attaining HbA1c goals (< 7%) compared to usual care[27]. However, this was a high risk of bias study, scoring ‘unclear’ across the six domains.

##### Diastolic blood pressure

Of 11 studies which reported diastolic blood pressure (DBP)[27,30,33–36,38,41,42,45,50,54], eight were included in meta-analysis[27,33,35,38,41,42,45,54]. A very small statistically significant pooled effect was found at follow-up (*d*=-0.123, *95%CI* = -0.22, -0.03, *n=*8) (p=0.008), whereby participants in the SMA group had lower DBP than those in usual care (see Figure 2b).

Of the three studies not included in the meta-analysis[30,36,50], one trial of SMAs for diabetes and hypertension reported that mean DBP was lower in the SMA group (78.3 mmHg) than in the usual care group (82.1 mmHg) at 12 months[50].

##### Systolic blood pressure

Of 13 trials reporting SBP[27,30,33,35,36,38,41,42,44–46,50,54], nine could be meta-analysed[27,33,35,38,41,42,45,46,54]. No statistically significant difference between SMAs and usual care was found for SBP at follow-up (*d*=-0.018, *95%CI* = -0.11, 0.08, *n=*9) (*p*=0.709), with lows level of heterogeneity observed (*I^2^ =* 3.8%) (see Figure 2c). Of the four trials not in the meta-analysis[30,36,44,50], two moderately robust studies reported statistically significant between group differences in SBP at follow-up[36,50], whereby the SMA group showed greater decreases in SBP compared to usual care.

No statistically significant effect of SMAs compared to usual care was found for other biomedical health outcomes including: total cholesterol, high density lipoprotein cholesterol, low-density lipoprotein cholesterol, triglycerides, weight, and BMI (see S6).

##### Trials with non-usual care comparators

Three trials had enhanced SMAs as their comparator[34,37,48], all of which found greater reductions in the enhanced SMA group compared to the standard SMA group [64,68,69]. Drake et al. (2018) reported that there significantly greater improvements in HbA1c (%) were observed in the PHP SMA group compared to the standard SMA group at follow-up[68]. Naik et al. (2011) found that HbA1c (%) was significantly lower in the SMA group than the traditional diabetes group education group immediately following the active interventions at three months, and the between group differences remained clinically and statistically significant at one year follow-up[64]. Yancy et al. (2020) found that the mean reduction in HbA1c (%) was significantly greater in the enhanced SMA group compared to the standard GMV group at 16 and 32 weeks. However, at 48 weeks, no between group differences in HbA1c (%) were observed[69].

Further, Drake et al., (2018) reported that participants in the PHP SMA group had lower DBP (*M*=86 mmHg) at 8 months follow-up compared to participants in the standard SMA group (*M*=79.8 mmHg)[34]. Yancy et al. (2020) reported patient weight loss in the SMAs with intensive weight management was comparable to weight loss amongst patients attending SMAs but statistical significance was unclear[48]. It is possible that other studies could not detect statistically significant differences between arms due to small sample sizes.

**Figure 2:** Forest plot for a) HbA1C(%), b) diastolic blood pressure, c) systolic blood pressure

#### Psychological and well-being measures

##### Quality of life

Six trials reported quality of life (QoL) outcomes[23,26,35,38,44,46] of which two trials reported significant between group differences[23,38]. One trial of SMAs for chronically ill patients with multiple LTCs found that participants in the SMA group (*M*=7.2, *SD*=1.8) reported significantly better QoL than the usual care group (*M*=6.3, *SD*=2.0) (*p*=.002) at 24 months[23]. Schillinger et al., (2008) measured QoL using the short form (SF)-12 instrument which comprised of mental health and physical health subscales. Improvements in SF-12 mental health was observed for SMA group compared to SMA (effect size=0.31, *p*=0.03) and usual care (effect size=0.18, *p*=0.2)[38]. However, this was considered as a high risk of bias study, with high/unclear judgements across four out of six domains.

##### Patient satisfaction

Four trials measured patient satisfaction[23,34,43,56]. Scott et al. (2004) reported significant differences at follow-up, with SMA patients reporting higher satisfaction with practitioner discussions compared to controls[23]. The other three studies found no between group differences.

##### Patient self-efficacy

Self-efficacy was measured in nine trials[23,27,36,38,44,47,50,54], of which five studies were included in meta-analysis[23,26,27,38,54]. No statistically significant effect was found (*d*=0.167, *95%CI* = -0.08, 0.41, *n=*5) (p=0.182) (See S7). High levels of heterogeneity were observed (*I^2^*=78.9%). Of the four other studies not included in meta-analysis[36,44,47,50], two reported that SMA patients had significant improvements in self-efficacy to manage diabetes compared to usual care[36,50].

No statistically significant effect of SMAs compared to usual care was found for other depression, the only other psychological and well-being measure identified (see S7).

##### Trials with non-usual care comparator

Drake et al. (2018) reported significant improvements in self-efficacy, as measured using the Diabetes Empowerment Scale, for the PHP SMA group compared to the standard SMA group[30]. Naik et al. (2011) did not find any differences in diabetes self-efficacy scores between the SMA group and the traditional diabetes group education group[37].

#### Healthcare service utilisation

##### Hospital admissions

Seven trials reported hospital admissions within six to 24 months[23,27,29,43,45,47,50] and three were included in a meta-analysis[23,27,45]. There was no difference between SMAs and usual care in terms of hospital admissions at follow-up (*d*=-0.016, *95%CI* = -0.38, 0.35, *n=*3; (*p*=0.931) (see Figure 3a). Substantial heterogeneity was observed (*I*^2^ = 71.1%).

None of the other four trials[29,43,47,50] reported significant between group differences for hospital admissions at follow-up.

##### Emergency department use

Of eight relevant trials[23,26,27,29,43,45,47,50], four were included in a meta-analysis[23,27,43,45]. No difference between SMAs and usual care was found for admissions to emergency departments at follow-up (*d*=-0.083, *95%CI* = -0.30, 0.13, *n=*4, *p*=0.453 (Figure 3b). Considerable heterogeneity was observed (*I*^2^ = 61.7%).

Of four trials not in the meta-analysis [26,29,47,50], only Edelman et al. (2010) reported significant between group differences in emergency department use favouring SMAs with 0.4 (*95%CI* = 0.20, 0.70) fewer emergency care visits than the usual care group over the 12-month study period [50].

##### Primary care visits

Four trials reported the number of primary care visits participants made during the study period[23,43,47,50]. Three were pooled in a meta-analysis[23,43,47] showing no statistically significant difference (*d*=0.034, *95%CI* = -0.09, 0.16, *n=*3, *p*=0.575) (see Figure 3c).

Edelman et al. (2010), which could not be included into the meta-analysis, reported that SMA participants had significantly fewer primary care visits than controls (5.3 vs. 6.2 per patient-year) at 12 months[50].

No statistically significant effect of SMAs compared to usual care were found for other behavioural outcomes including medication adherence and physical activity (see S8)

**Figure 3:** Forest plot for a) hospital admissions, b) emergency department use, c) primary care visits

#### Cost and resource use

Few studies reported the costs involved in the delivery of the SMA and those that did were unclear about cost parameters (i.e whether scheduling and preparation time was included or not) or how the costs were attributed. One trial of diabetes SMAs reported that overall costs per patient were higher in SMAs than those in usual care group for the study period of six months[29]. However, another trial found no significant difference between SMA and usual care in terms of total costs incurred during the 24 months study period, but showed a positive effect of the SMA at 13 months post-study where cost decreased by 6% for the SMA but increased by 13% for usual care *p*<0.01[43]. The SMA trial for osteoporosis reported that the costs incurred during the study period were lower for the SMA group compared to control groups[56]. A trial of chronic condition SMAs (reported that total costs incurred by the SMA group were lower than the usual care group[23].

## DISCUSSION

Our systematic review identified 23 unique RCTs comparing SMAs for one or more LTCs to usual care or an enhanced SMA. We found that SMAs significantly improved diastolic blood pressure for diabetes patients. In line with the findings of previous reviews[15], no harm was observed for the use of SMAs across these outcomes and there was not enough evidence of an effect on healthcare service use compared to usual care. This indicates that whilst SMAs may not be superior to usual care in terms of most health outcomes or reducing demand on services, they do not appear to increase demand at least in the short-term. Evidence reporting costs is too heterogeneous to draw firm conclusions.

### Comparison with previous literature

Like previous reviews of SMAs for LTCs[4,15,70], more than half of the included RCTs included patients with diabetes, and as such the most commonly reported outcome measure was HbA1C. However, unlike previous reviews[71,72], we did not observe any significant improvements in HbA1c. This may be because previous reviews included trials in secondary care. Our meta-analysis showed that SMA participants had lower DBP compared to patients who received usual care. [54]

Previous systematic reviews have been inconclusive with regards to the impact of SMAs on healthcare utilisation. Edelman et al.’s (2012) review[72] of SMAs for patients with chronic medical conditions in older adults found a lower pattern of healthcare utilisation, whilst Booth et al. (2015)[4] reported a mixed pattern of changes. Our meta-analyses show that SMAs do not differ from usual care in terms of healthcare utilisation. There is no evidence in the present data to suggest that patients compensate for a lack of privacy by returning to primary care or that they risk hospitalisation because issues are not adequately addressed during the SMA session(s). However, it should be noted that the key source of bias across the included studies was the lack of blinding of participants and personnel. Therefore, possible selection bias may result in recruitment of SMA participants with less concern about sharing their personal/medical information.

In comparison to biomedical outcomes and psychological outcomes, healthcare service use and costs and resource use and other behavioural outcomes were less frequently reported by studies. This echoes the findings of Edelman et al. (2015)[46] which found there to be limited data on key patient-centred outcomes such as patient satisfaction. Behavioural outcomes such as medication adherence are important across many LTCs and are key to understanding how patients are self-managing their conditions. In line with Kelly et al.’s (2017)[12] recommendation, future studies should report outcome effectiveness measures that are common or comparable across different LTCs such as physical activity, self-efficacy, medication adherence, and quality of life. It would be advantageous to agree a Core Outcome Set (COS), consisting of a standardised group of outcomes, to be reported by all SMA trials. This can help with future evaluations of SMAs through reducing heterogeneity and facilitating meta-analysis and ensuring that outcome measures are relevant to key stakeholders[73].

### Strengths and limitations

Although previous reviews have explored the effectiveness of SMAs in improving health outcomes, this review provides a focus on primary care which is key to managing LTCs. We found ten additional trials with 1160 participants since the comprehensive work by Booth et al. (2015)[4] indicating a rapidly growing field. We used robust methods whereby our search strategy was developed with input from Information Specialists through an iterative process and key stages of the review (including screening, data extraction, and quality appraisal) were undertaken independently by two reviewers. We included studies regardless of type of LTC so that we were able to summarise all the available evidence on effectiveness of SMAs for LTCs in primary care in one analysis. However, evidence of an effect was determined by *p*=<.0.5 in the papers. This assumes that studies were adequately powered, which may not be the case, particularly for some of the secondary outcomes of the included studies.

### Limitations of evidence base and implications for future research

Despite using a form of delivery framework to extract relevant study information[17], some important contextual factors, such characteristics of the healthcare professionals delivering the SMA, may not have been captured as this information was missing from the authors’ descriptions of the SMAs. Similar components may be described differently by different authors or, conversely, similar descriptions are used to describe different components. Using standardised taxonomies for describing form of delivery and intervention content when designing the intervention/SMA content could help to identify important behavioural components and key implementation processes that contribute to intervention effectiveness, allowing for replication. However, for this to be possible, it is also important that interventionists clearly specify which target behaviours (e.g. to increase physical activity) the SMAs aim to change. None of the included studies included measures of fidelity which is also important for determining whether the session(s) are delivered as intended to achieve optimum effects[74]. Further, theoretical underpinnings were lacking in the included SMA interventions, making it difficult to identify ‘mechanisms of action’ through which interventions bring about change[75]. Future SMAs interventions should be theory-based and be explicit in reporting its theoretical underpinnings.

Where multiple healthcare professionals are involved in the SMA, their key role and purpose in the SMA were rarely clearly defined. There was also limited reporting on the composition of the SMA groups across some of the included studies (i.e. how patients were selected for recruitment and size). Therefore, it is unclear which groups of patients might benefit from attending the same SMAs together and what implications SMAs may have for intervention-generated health inequalities. One third of the included studies were conducted on US veterans and one third of studies have involved participants from low-income/uninsured population groups. Generalisability of these groups to other healthcare settings in other countries is unclear. There needs to be further examination into how SMAs have been implemented into typical NHS practice. It was envisaged that “they are not an addition to one-to-one appointments – they replace them.” (p.65, Clay & Stern, 2015). However, there is anecdotal evidence that SMAs are being used in addition to usual care models of chronic disease management rather than as replacements. Further investigations into SMAs for patients with one or more LTCs is required, including a wider variety of LTCs (such as asthma and chronic obstructive pulmonary disease) and with more diverse population groups. For example, including low-income and disadvantaged groups in other countries, including the UK.

### Conclusions

This review is the first to examine the effects of SMAs delivered in primary care on health outcomes and healthcare service use in patients with one or more LTCs. Our review suggests that SMAs are unlikely to result in less favourable outcomes to patients with LTCs compared to usual care. To identify key intervention components that contribute to effectiveness, future studies will benefit from using standardised taxonomies to report intervention content. The use of an evaluation framework, with a core outcome set, is recommended to improve evidence in this field.

## AUTHOR CONTRIBUTIONS

EK and FS designed the study. MYT, FG, CR, and FB developed the search strategy and undertook the searches. FG and MYT carried out the screening, data extraction, and analysis. All authors contributed to the interpretation. MYT and FG wrote the manuscript that all authors contributed to and approved.

## Data Availability

All data produced in the present work are contained in the manuscript.

## ACKNOWLEDGMENTS

We would like to thank the PRU BehSci Public Patient Involvement group for providing their patient perspective about outcome measures of interests. Professor Kaner is supported via an NIHR Senior Investigator award.

## ETHICS STATEMENT

Ethical approval was not required due to the present study being a systematic review.

## AVAILABILITY OF DATA

As this study is a systematic review, all data reported has been previously published and is in the public domain.

## SUPPLEMENTARY MATERIAL

PROSPERO PROTOCOL

PRISMA checklist

Supplementary File 1- Search strategy for Ovid MEDLINE

Supplementary File 2- Data Extraction Form

Supplementary File 3- List of included peer-reviewed papers

Supplementary File 4- Outcomes reported by studies

Supplementary File 5- Quality assessment of included studies with sensitivity analyses

Supplementary File 6- Additional biomedical measures examined with forest plots

Supplementary File 7- Additional psychological and well-being measures examined with forest plots

Supplementary File 8- Additional behavioural outcomes examined with forest plots

Supplementary File 9- SMA delivery, dose and components

## REFERENCES

1 Clay H, Stern R. Making time in general practice: freeing GP capacity by reducing bureaucracy and avoidable consultations, managing the interface with hospitals and exploring new ways of working. 2015.

2 Hayhoe B, Verma A, Kumar S. Shared medical appointments. BMJ (Online) 2017;358:10–1. doi:10.1136/bmj.j4034

3 Jones T, Darzi A, Egger G, et al. Process and Systems: A systems approach to embedding group consultations in the NHS. Future Healthcare Journal 2019;6:8–16. doi:10.7861/futurehosp.6-1-8

4 Booth A, Cantrell A, Preston L, et al. What is the evidence for the effectiveness, appropriateness and feasibility of group clinics for patients with chronic conditions? A systematic review. Health Services and Delivery Research 2015;3:1–194. doi:10.3310/hsdr03460

5 Clay H, Stern R. Making Time in General Practice. 2015. https://www.primarycarefoundation.co.uk/images/PrimaryCareFoundation/Downloading_Reports/PCF_Press_Releases/Making-Time-in_General_Practice_FULL_REPORT_28_10_15.pdf

6 Wadsworth KH, Archibald TG, Payne AE, et al. Shared medical appointments and patient-centered experience: A mixed-methods systematic review. BMC Family Practice 2019;20:1–13. doi:10.1186/s12875-019-0972-1

7 Graham F, Tang MY, Jackson K, et al. Barriers and facilitators to implementation of shared medical appointments in primary care for the management of long-term conditions: a systematic review and synthesis of qualitative studies. BMJ Open 2021;11:e046842. doi:10.1136/bmjopen-2020-046842

8 Edelman D, Gierisch JM, McDuffie JR, et al. Shared Medical Appointments for Patients with Diabetes Mellitus: A Systematic Review. Journal of General Internal Medicine 2015;30:99–106. doi:10.1007/s11606-014-2978-7

9 Booth A, Cantrell A, Preston L, et al. What is the evidence for the effectiveness, appropriateness and feasibility of group clinics for patients with chronic conditions? A systematic review. Health Services and Delivery Research 2015;3:1–194. doi:10.3310/hsdr03460

10 Higgins J, Thomas J, Chandler J, et al. Systematic Reviews of Interventions. In: Deeks J, Higgins J, Altman D, eds. Cochrane Handbook for Systematic Reviews of Interventions version 6.2 (updated February 2021). 2021.

11 Page MJ, McKenzie JE, Bossuyt PM, et al. The PRISMA 2020 statement: An updated guideline for reporting systematic reviews. The BMJ 2021;372. doi:10.1136/bmj.n71

12 Michie S, Richardson M, Johnston M, et al. The behavior change technique taxonomy (v1) of 93 hierarchically clustered techniques: Building an international consensus for the reporting of behavior change interventions. Annals of Behavioral Medicine 2013;46:81–95. doi:10.1007/s12160-013-9486-6

13 Booth A, Cantrell A, Preston L, et al. What is the evidence for the effectiveness, appropriateness and feasibility of group clinics for patients with chronic conditions? A systematic review. Health Services and Delivery Research 2015;3:1–194. doi:10.3310/hsdr03460

14 Kirsh SR, Aron DC, Johnson KD, et al. A realist review of shared medical appointments: How, for whom, and under what circumstances do they work? BMC Health Services Research 2017;17:1–13. doi:10.1186/s12913-017-2064-z

15 Kelly F, Liska C, Morash R, et al. Shared medical appointments for patients with a nondiabetic physical chronic illness: A systematic review. Chronic Illness 2019;15:3– 26. doi:10.1177/1742395317731608

16 Veritas Health Innovation. Covidence systematic review software. 2021.

17 Dombrowski SU, O’Carroll RE, Williams B. Form of delivery as a key ‘active ingredient’ in behaviour change interventions. British Journal of Health Psychology 2016;21:733–40. doi:10.1111/bjhp.12203

18 Higgins JPT, Altman DG, Gøtzsche PC, et al. The Cochrane Collaboration’s tool for assessing risk of bias in randomised trials. BMJ (Online) 2011;343:1–9. doi:10.1136/bmj.d5928

19 Centre for Reviews and Dissemination. CRD’s guidance for undertaking reviews in health care. York: 2009.

20 Valentine JC, Pigott TD, Rothstein HR. How many studies do you need? A primer on statistical power for meta-analysis. Journal of Educational and Behavioral Statistics 2010;35:215–47. doi:10.3102/1076998609346961

21 Deeks J, Higgins J, Altman D. Analysing data and undertaking meta-analyses. In: Higgins J, Thomas J, Chandler J, et al., eds. Cochrane Handbook for Systematic Reviews of Interventions. Cochrane 2021.

22 Popay J, Roberts H, Sowden A, et al. Guidance on the Conduct of Narrative Synthesis in Systematic Reviews A Product from the ESRC Methods Programme Peninsula Medical School, Universities of Exeter and Plymouth. 2006.

23 Scott JC, Conner DA, Venohr I, et al. Effectiveness of a group outpatient visit model for chronically ill older health maintenance organization members: A 2-year randomized trial of the Cooperative Health Care Clinic. J Am Geriatr Soc 2004;52:1463–70. doi:10.1111/j.1532-5415.2004.52408.x

24 Coleman EA, Eilertsen TB, Kramer AM, et al. Reducing emergency visits in older adults with chronic illness. A randomized, controlled trial of group visits. Eff Clin Pract 2001;4:49–57.

25 Gardiner P, Lestoquoy AS, Gergen-Barnett K, et al. Design of the integrative medical group visits randomized control trial for underserved patients with chronic pain and depression. Contemporary Clinical Trials 2017;54:25–35. doi:10.1016/j.cct.2016.12.013

26 Gardiner P, Luo M, D’Amico S, et al. Effectiveness of integrative medicine group visits in chronic pain and depressive symptoms: A randomized controlled trial. PLoS ONE 2019;14:1–20. doi:10.1371/journal.pone.0225540

27 Berry DC, Williams W, Hall EG, et al. Imbedding Interdisciplinary Diabetes Group Visits Into a Community-Based Medical Setting. The Diabetes Educator 2016;42:96– 107. doi:10.1177/0145721715620022

28 Clancy DE, Cope DW, Magruder K, et al. Evaluating group visits in an uninsured or inadequately insured patient population with uncontrolled type 2 diabetes. Diabetes Educ 2003;29:292–203. doi:10.1177/0145721707299266

29 Clancy DE, Cope DW, Magruder KM, et al. Evaluating concordance to American Diabetes Association standards of care for type 2 diabetes through group visits in an uninsured or inadequately insured patient population. Diabetes Care 2003;26:2032–6. doi:10.2337/diacare.26.7.2032

30 Clancy DE, Huang P, Okonofua E, et al. Group visits: Promoting adherence to diabetes guidelines. Journal of General Internal Medicine 2007;22:620–4. doi:10.1007/s11606-007-0150-3

31 Clancy DE, Yeager DE, Huang P, et al. Further evaluating the acceptability of group visits in an uninsured or inadequately insured patient population with uncontrolled type 2 diabetes. Diabetes Educ 2007;33:309–14. doi:10.1177/0145721707299266

32 Clancy DE, Dismuke CE, Magruder KM, et al. Do Diabetes Group Visits Lead to Lower Medical Care Charges? American Journal of Managed Care 2008;14:39–44.

33 Cole RE, Boyer KM, Spanbauer SM, et al. Effectiveness of Prediabetes Nutrition Shared Medical Appointments: Prevention of Diabetes. The Diabetes Educator 2013;39:344–53. doi:10.1177/0145721713484812

34 Drake C, Meade C, Hull SK, et al. Integration of Personalized Health Planning and Shared Medical Appointments for Patients with Type 2 Diabetes Mellitus. Southern Medical Journal 2018;111:674–82. doi:10.14423/SMJ.0000000000000892

35 Ee C, de Courten B, Avard N, et al. Shared Medical Appointments and Mindfulness for Type 2 Diabetes—A Mixed-Methods Feasibility Study. Frontiers in Endocrinology 2020;11:1–16. doi:10.3389/fendo.2020.570777

36 Liu S, Bi A, Fu D, et al. Effectiveness of using group visit model to support diabetes patient self-management in rural communities of Shanghai: A randomized controlled trial. BMC Public Health 2012;12:1. doi:10.1186/1471-2458-12-1043

37 Naik AD, Palmer N, Petersen NJ, et al. Comparative effectiveness of goal setting in diabetes mellitus group clinics: Randomized clinical trial. Archives of Internal Medicine 2011;171:453–9. doi:10.1001/archinternmed.2011.70

38 Schillinger D, Hammer H, Wang F, et al. Seeing in 3-D: Examining the reach of diabetes self-management support strategies in a public health care system. Health Education and Behavior 2008;35:664–82. doi:10.1177/1090198106296772

39 Schillinger D, Handley M, Wang F, et al. Effects of self-management support on structure, process, and outcomes among vulnerable patients with diabetes: a 3-arm randomized trial. Diabetes Care 2009;32. doi:10.2337/dc08-0787.Clinical

40 Wallace A, Perkhounkova Y, Tseng H, et al. Influence of patient characteristics on assessment of diabetes self-management support. Nursing Research 2013;62:106– 14. doi:10.1097/NNR.0b013e3182843b77

41 Vaughan EM, Johnston CA, Cardenas VJ, et al. Integrating CHWs as Part of the Team Leading Diabetes Group Visits: A Randomized Controlled Feasibility Study. Diabetes Educator 2017;43:589–99. doi:10.1177/0145721717737742

42 Vaughan EM, Hyman DJ, Naik AD, et al. A Telehealth-supported, Integrated care with CHWs, and MEdication-access (TIME) Program for Diabetes Improves HbA1c: a Randomized Clinical Trial. Journal of General Internal Medicine Published Online First: 2020. doi:10.1007/s11606-020-06017-4

43 Wagner EH, Grothaus LC, Sandhu N, et al. Chronic care clinics for diabetes in primary care: A system-wide randomized trial. Diabetes Care 2001;24:695–700. doi:10.2337/diacare.24.4.695

44 Cohen LB, Taveira TH, Khatana SAM, et al. Pharmacist-Led Shared Medical Appointments for Multiple Cardiovascular Risk Reduction in Patients With Type 2 Diabetes. The Diabetes Educator 2011;37:801–12. doi:10.1177/0145721711423980

45 Taveira TH, Wu WC. Interventions to maintain cardiac risk control after discharge from a cardiovascular risk reduction clinic: A randomized controlled trial. Diabetes Research and Clinical Practice 2014;105:327–35. doi:10.1016/j.diabres.2014.05.013

46 Wu WC, Taveira TH, Jeffery S, et al. Costs and effectiveness of pharmacist-led group medical visits for type-2 diabetes: A multi-center randomized controlled trial. PLoS ONE 2018;13:1–14. doi:10.1371/journal.pone.0195898

47 Taveira TH, Dooley AG, Cohen LB, et al. Pharmacist-Led Group Medical Appointments for the Management of Type 2 Diabetes with Comorbid Depression in Older Adults. Annals of Pharmacotherapy 2011;45:1346–55. doi:10.1345/aph.1Q212

48 Yancy WS, Crowley MJ, Dar MS, et al. Comparison of Group Medical Visits Combined with Intensive Weight Management vs Group Medical Visits Alone for Glycemia in Patients with Type 2 Diabetes: A Noninferiority Randomized Clinical Trial. JAMA Internal Medicine 2020;180:70–9. doi:10.1001/jamainternmed.2019.4802

49 Crowley MJ, Edelman D, Voils CI, et al. Jump starting shared medical appointments for diabetes with weight management: Rationale and design of a randomized controlled trial. Contemporary Clinical Trials 2017;58:1–12. doi:10.1016/j.cct.2017.04.004

50 Edelman D, Fredrickson SK, Melnyk SD, et al. Medical Clinics Versus Usual Care for Patients With Both Diabetes. Annals of Internal Medicine 2010;152:689–96.

51 Crowley MJ, Melnyk SD, Coffman CJ, et al. Impact of baseline insulin regimen on glycemic response to a group medical clinic intervention. Diabetes Care 2013;36:1954–60. doi:10.2337/dc12-1905

52 Crowley MJ, Melnyk SD, Ostroff JL, et al. Can group medical clinics improve lipid management in diabetes? American Journal of Medicine 2014;127:145–51. doi:10.1016/j.amjmed.2013.09.027

53 Eisenberg A, Crowley MJ, Coffman C, et al. Effect of a group medical clinic for veterans with diabetes on body mass index. Chronic Illness 2019;15:187–96. doi:10.1177/1742395317753885

54 Gao J, Yang L, Junming D, et al. Evaluation of group visits for Chinese hypertensives based on primary health care center. Asia-Pacific Journal of Public Health 2015;27:NP350–60. doi:10.1177/1010539512442566

55 Simon B, Sawicki PT. Quality improvement in chronic care delivery for patients with arterial hypertension through Group Medical Visits: Patient acceptance and attendance in the German pilot project. Quality in Primary Care 2015;23:106–112 7p.

56 Baqir W, Gray WK, Blair A, et al. Osteoporosis group consultations are as effective as usual care: Results from a non-inferiority randomized trial. Lifestyle Medicine 2020;1:1–9. doi:10.1002/lim2.3

57 Gardiner P, Luo M, D’Amico S, et al. Effectiveness of integrative medicine group visits in chronic pain and depressive symptoms: A randomized controlled trial. PLoS ONE 2019;14:1–20. doi:10.1371/journal.pone.0225540

58 Berry DC, Williams W, Hall EG, et al. Imbedding Interdisciplinary Diabetes Group Visits Into a Community-Based Medical Setting. The Diabetes Educator 2016;42:96–107. doi:10.1177/0145721715620022

59 Clancy DE, Cope DW, Magruder K, et al. Evaluating group visits in an uninsured or inadequately insured patient population with uncontrolled type 2 diabetes. Diabetes Educ 2003;29:292–203. doi:10.1177/0145721707299266

60 Clancy DE, Huang P, Okonofua E, et al. Group visits: Promoting adherence to diabetes guidelines. Journal of General Internal Medicine 2007;22:620–4. doi:10.1007/s11606-007-0150-3

61 Ee C, de Courten B, Avard N, et al. Shared Medical Appointments and Mindfulness for Type 2 Diabetes—A Mixed-Methods Feasibility Study. Frontiers in Endocrinology 2020;11:1–16. doi:10.3389/fendo.2020.570777

62 Vaughan EM, Hyman DJ, Naik AD, et al. A Telehealth-supported, Integrated care with CHWs, and MEdication-access (TIME) Program for Diabetes Improves HbA1c: a Randomized Clinical Trial. Journal of General Internal Medicine Published Online First: 2020. doi:10.1007/s11606-020-06017-4

63 Baqir W, Gray WK, Blair A, et al. Osteoporosis group consultations are as effective as usual care: Results from a non-inferiority randomized trial. Lifestyle Medicine 2020;1:1–9. doi:10.1002/lim2.3

64 Naik AD, Palmer N, Petersen NJ, et al. Comparative effectiveness of goal setting in diabetes mellitus group clinics: Randomized clinical trial. Archives of Internal Medicine 2011;171:453–9. doi:10.1001/archinternmed.2011.70

65 Liu S, Bi A, Fu D, et al. Effectiveness of using group visit model to support diabetes patient self-management in rural communities of Shanghai: A randomized controlled trial. BMC Public Health 2012;12:1. doi:10.1186/1471-2458-12-1043

66 Crowley MJ, Melnyk SD, Coffman CJ, et al. Impact of baseline insulin regimen on glycemic response to a group medical clinic intervention. Diabetes Care 2013;36:1954–60. doi:10.2337/dc12-1905

67 Taveira TH, Wu WC. Interventions to maintain cardiac risk control after discharge from a cardiovascular risk reduction clinic: A randomized controlled trial. Diabetes Research and Clinical Practice 2014;105:327–35. doi:10.1016/j.diabres.2014.05.013

68 Drake C, Meade C, Hull SK, et al. Integration of Personalized Health Planning and Shared Medical Appointments for Patients with Type 2 Diabetes Mellitus. Southern Medical Journal 2018;111:674–82. doi:10.14423/SMJ.0000000000000892

69 Yancy WS, Crowley MJ, Dar MS, et al. Comparison of Group Medical Visits Combined with Intensive Weight Management vs Group Medical Visits Alone for Glycemia in Patients with Type 2 Diabetes: A Noninferiority Randomized Clinical Trial. JAMA Internal Medicine 2020;180:70–9. doi:10.1001/jamainternmed.2019.4802

70 Edelman D, McDuffie JR, Oddone E, et al. Shared Medical Appointments for Chronic Medical ConditionsL: A Systematic Review. VA Evidence-based Synthesis Program Reports 2012.

71 Housden L, Wong ST, Dawes M. Effectiveness of group medical visits for improving diabetes care: a systematic review and meta-analysis. Canadian Medical Association Journal 2013;185:E635–44. doi:10.1503/cmaj.130053

72 Edelman D, Gierisch JM, McDuffie JR, et al. Shared Medical Appointments for Patients with Diabetes Mellitus: A Systematic Review. Journal of General Internal Medicine 2015;30:99–106. doi:10.1007/s11606-014-2978-7

73 Webbe J, Sinha I, Gale C. Core Outcome Sets. Archives of Disease in Childhood: Education and Practice Edition 2018;103:163–6. doi:10.1136/archdischild-2016-312117

74 Bellg AJ, Resnick B, Minicucci DS, et al. Enhancing treatment fidelity in health behavior change studies: Best practices and recommendations from the NIH Behavior Change Consortium. Health Psychology 2004;23:443–51. doi:10.1037/0278-6133.23.5.443

75 Carey RN, Connell LE, Johnston M, et al. Behavior Change Techniques and Their Mechanisms of Action: A Synthesis of Links Described in Published Intervention Literature. Annals of Behavioral Medicine 2018;53:693–707. doi:10.1093/abm/kay078

