## Supplementary materials for "Effectiveness of shared medical appointments delivered in primary care for improving health outcomes in patients with long-term conditions: a systematic review of randomised controlled trials"

### S1 – Search strategy for Ovid MEDLINE

**Ovid MEDLINE(R) and Epub Ahead of Print, In-Process & Other Non-Indexed Citations, Daily and Versions(R)** 1946 to September 09, 2020

| # | Search Term | Search results |
|---|---|---|
| 1 | (group adj1 visit*).tw. | 719 |
| 2 | (group adj3 appointment*).tw. | 232 |
| 3 | (group medical adj2 visit*).tw. | 83 |
| 4 | (group medical adj2 clinic*).tw. | 14 |
| 5 | (group medical adj2 appointment*).tw. | 28 |
| 6 | (group medical adj2 care).tw. | 11 |
| 7 | (group medical adj2 meeting*).tw. | 0 |
| 8 | (group adj3 consultation*).tw. | 724 |
| 9 | (shared medical adj2 appointment*).tw. | 159 |
| 10 | (shared medical adj2 visit*).tw. | 10 |
| 11 | (cluster adj3 visit*).tw. | 51 |
| 12 | (group medical adj2 consultation*).tw. | 5 |
| 13 | (chronic care adj2 clinic*).tw. | 42 |
| 14 | exp Shared Medical Appointments/ | 24 |
| 15 | (group clinic*).tw. | 3085 |
| 16 | /OR 1-15 | 5041 |
| 17 | Limit 16 to yr= ”2013-Current” | 2500 |

### S2 – Data Extraction Form

| Section on Covidence | Field | Notes | Codes |
|---|---|---|---|
| ***Identification*** | *Country* | See codes | 1- Australia<br>2- Canada<br>3- China<br>4- Netherlands<br>5- United Kingdom<br>6- United States<br>7- Other |
| | *Primary care setting* (the default 'Setting' item on Covidence) | See codes | 1- GP surgery<br>2- Community health centre<br>3- Other (specify as free text)<br>4- Unclear<br>888- Not reported |
| | *Comments* | Free text (if applicable) | |
| | *Setting (urban/rural)* | Free text – e.g., in city setting | |
| ***Methods*** | *Design* | Choose from drop-down list options | |
| | *Aim of the study* | Free text | |
| | *Primary outcome* | Free text - describe reported primary outcome | |
| | *Secondary outcome(s)* | Free text - describe any reported secondary outcome(s) | |
| | *Statistical analysis* | Free text - brief description of statistical analysis, including any details on intention-to-treat and adjustment for clustering (where appropriate). Note any problems/errors. | |
| | *Timepoints reported* | Free text - list all reported, including baseline, in months since randomisation (for compliance with target behaviour) | |
| ***Population*** | *Inclusion criteria* | Free text | |
| | *Exclusion criteria* | Free text | |
| | *Group differences* | Free text | |
| | *Clinical condition(s) for which SMA was held for* | See codes | 1- Diabetes<br>2- Osteoporosis only<br>3- Chronic pain only<br>4- Chronic neuromuscular disease only<br>5- Multimorbidity<br>6- Other (specify as free text) |
| | *Description of multimorbidity (where relevant)* | Free text | |
| | *Method(s) of recruitment of patients* | Free text | |
| | *Number of clusters (if applicable)* | Free text – only for cluster randomised controlled trials | |
| | *Subgroups measured and/or reported* | Free text | |
| | *Total number randomised (or total population if cluster randomised)* | Free text | |
| | *Were disadvantaged patient (groups) targeted/recruited?* | See codes | 0- No<br>1- Homelessness<br>2- Substance misuse |

| | | | |
|---|---|---|---|
| | | | 3- Offending behavioural issues<br>4- Mental ill-health<br>5- Other (specify as free text)<br>6- Unclear |
| | *Withdrawals and exclusions (after randomisation)* | Free text (include details on dropouts/non-attendance at sessions) | |
| | *Baseline characteristics – age (mean)* | Free text | |
| | *Baseline characteristics – age (standard deviation)* | Free text | |
| | *Baseline characteristics – number of male participants* | Free text | |
| | *Baseline characteristics – number of female participants* | Free text | |
| | *Baseline characteristics – number of White participants* | Free text | |
| | *Baseline characteristics – number of non-White participants* | Free text | |
| | *Baseline characteristics – education (number)* | Free text | |
| | *Baseline characteristics – employment (number)* | Free text | |
| ***Interventions*** | *Intervention/control name* | Free text – name given by authors | |
| | *Number of participants randomised to group* | Free text | |
| | *Number of clusters randomised to group (CRCT only)* | Free text – only relevant for cluster randomised controlled trials, (777 if NA) | |
| | *Authors' description of intervention (e.g., SMA, GV, GC)* | Copy and paste *all* details given by the authors about the intervention. Please include details from any of the study papers.<br><br>Flag or paste link(s) to supplementary material(s), where appropriate | |
| | *Description of co-interventions (e.g., SMA + Peer 2 peer support)* | Free text | |
| | *Provider characteristics* | Free text – e.g., male; female; age; ethnicity<br><br>*Involved in delivery of the SMAs (including facilitators)*<br><br>*Involved in set-up/organisation of the SMAs* | |
| | *Professional background (delivery of the SMAs, including facilitators)* | See codes – list all that apply | 1- GP<br>2- Other doctor<br>3- Nurse<br>4- Dietician/nutritionist<br>5- Pharmacist<br>6- Psychologist |

| | | | |
|---|---|---|---|
| | | | 7- Physiotherapist<br>8- Counsellor<br>9- Social worker<br>10- Other allied healthcare professional (specify as free text)<br>11- Other non-healthcare professional (specify as free text)<br>12- Unclear<br>888- Not reported |
| | *Professional background (organisation/set-up of the SMAs)* | Free text | |
| | *Professional experience* | Free text – e.g., 15 years practicing nurse, Masters level psychology student, registered dietician for 8 years<br><br>*Involved in delivery of the SMAs (including facilitators)*<br><br>*Involved in set-up/organisation of the SMAs* | |
| | *Training in intervention facilitation* | Free text - 2-hr training session; half day workshop; online training module | |
| | *Training in intervention delivery* | Free text – e.g., Communication skills training; group facilitation training, cognitive behavioural skills training | |
| | *Intervention relevant competence* | Free text – e.g., Competence Certified health trainer, certificate in counselling | |
| | *Continuity of providers who deliver the SMA sessions* | Free text – e.g., Same provider; different providers for different topics; mix of same and different providers | |
| | *Mode of delivery* | See codes | 1- Face-to-face<br>2- Teleconferencing<br>3- Face-to-face and teleconferencing<br>4- Other<br>5- Unclear<br>888- Not reported |
| | *Delivery channel* | Free text – e.g., Personal, self-help, mobile phone application (app), text message (SMS), telephone, email, CD-ROM, videoconferencing, podcast | |
| | *Delivery route* | See codes | 1- Audio<br>2- Text<br>3- Picture<br>4- Experiential<br>5- Unclear<br>888- Not reported |
| | *Participants' materials* | Free text – e.g., Leaflet, booklet, book, webpage, app, device<br>(e.g., pedometer), certificate, money, voucher | |
| | *Providers' materials* | Free text – e.g., Session manual; pop-up reminders, self-monitoring sheets | |
| | *Intervention materials* | Free text - e.g., Eligibility forms, questionnaires, sign in forms | |
| | *Venue of SMA delivery* | See codes | 1- GP surgery |

| | | | |
|---|---|---|---|
| | | | 2- Other hired venue (specify as free text)<br>3- Mixed<br>4- Unclear<br>888- Not reported |
| | *Duration of intervention (entire SMA programme)* | Free text | |
| | *Number of SMA sessions* | Free text | |
| | *Duration of SMA sessions* | Free text | |
| | *Frequency of SMA sessions* | Free text | |
| | *Spacing of SMA sessions* | Free text | |
| | *Contact form* | See codes | 1- Scheduled<br>2- Random<br>3- Proactive<br>4- Reactive<br>5- Unclear<br>888- Not reported |
| | *Intervention variation* | See codes | 1- None<br>2- One size for all<br>3- Personalised<br>4- Titrated<br>5- Adapted<br>6- Unclear<br>7- Other<br>888- Not reported |
| | *Tailoring source* | Free text – e.g. self-tailored, theory tailored, practitioner tailored. | |
| | *Standardisation* | Free text – e.g. an intervention manual is followed, automated, semi-automated, personal. | |
| | *Delivery style* | Free text – e.g. asset-based, patient-centred, authoritarian. | |
| | *Communication style* | Free text – e.g. patient led, practitioner led, narrative. | |
| | *Communication techniques* | Free text – e.g. listening, questioning, reflecting, pauses. | |
| | *Visual style* | Free text – e.g. logo, branding, colour scheme. | |
| | *Complexity* | Free text – e.g. reading level, layout, depth of information. | |
| | *Theoretical underpinnings* | See codes | 0- No<br>1- Yes (specify as free text)<br>2- Unclear |
| | *Costs reported/funding secured* | Free text | |
| | *Healthcare finance context- health insurance/free at point of use* | Free text | |

| ***Outcomes (e.g. HbA1C)*** | *Arm* | *Pre-intervention/baseline* | | | *Post-intervention* | | |
|---|---|---|---|---|---|---|---|
| | | Mean | Standard deviation | Sample size | Mean | Standard deviation | Sample size |
| | Intervention | | | | | | |
| | Control | | | | | | |

888 = Not reported; 777 = Not applicable; 999 = Missing data

**S3 – List of included peer-reviewed papers**

| Trial No. | Author & date | Paper title |
|---|---|---|
| *1* | *Baqir 2020* | Osteoporosis group consultations are as effective as usual care: Results from a non-inferiority randomized trial |
| 2 | *Berry 2016* | Imbedding Interdisciplinary Diabetes Group Visits Into a Community-Based Medical Setting |
| *3* | *Clancy 2003a* | Evaluating group visits in an uninsured or inadequately insured patient population with uncontrolled diabetes |
| *3* | *Clancy 2003b* | Evaluating Concordance to American Diabetes Association Standards of Care for Type 2 Diabetes Through Group Visits in an Uninsured or Inadequately Insured Patient Population |
| *4* | *Clancy 2007a* | Group Visits: Promoting Adherence to Diabetes Guidelines |
| 4 | Clancy 2007b | Further Evaluating the Acceptability of Group Visits in an Uninsured or Inadequately Insured Patient Population With Uncontrolled Type 2 Diabetes |
| *4* | *Clancy 2008* | Do Diabetes Group Visits Lead to Lower Medical Care Charges? |
| *5* | *Cohen 2011* | Pharmacist-Led Shared Medical Appointments for Multiple Cardiovascular Risk Reduction in Patients With Type 2 Diabetes |
| *6* | *Cole 2013* | Effectiveness of Prediabetes Nutrition Shared Medical Appointments |
| *7* | *Coleman 2001* | Reducing Emergency Visits in Older Adults with Chronic Illness. A Randomized, Controlled Trial of Group Visits |
| *7* | *Scott 2004* | Effectiveness of a Group Outpatient Visit Model for Chronically Ill Older Health Maintenance Organization Members: A 2-Year Randomized Trial of the Cooperative Health Care Clinic |
| 8 | Crowley (2013) | Impact of Baseline Insulin Regimen on Glycemic Response to a Group Medical Clinic Intervention |
| 8 | Crowley (2014) | Can Group Medical Clinics Improve Lipid Management in Diabetes? |
| 8 | Edelman (2010) | Medical Clinics Versus Usual Care for Patients With Both Diabetes and Hypertension: a randomised trial |
| *8* | *Eisenberg (2019)* | Effect of a group medical clinic for veterans with diabetes on body mass index |
| *9* | *Crowley (2017)* | Jump starting shared medical appointments for diabetes with weight management: Rationale and design of a randomized controlled trial |
| *9* | *Yancy (2020)* | Comparison of Group Medical Visits Combined With Intensive Weight Management vs Group Medical Visits Alone for Glycemia in Patients With Type 2 Diabetes A Noninferiority Randomized Clinical Trial |
| *10* | *Drake 2018* | Integration of Personalized Health Planning and Shared Medical Appointments for Patients with Type 2 Diabetes Mellitus |
| *11* | *Ee (2020)* | Shared Medical Appointments and Mindfulness for Type 2 Diabetes—A Mixed-Methods Feasibility Study |
| *12* | *Gao (2015)* | Evaluation of Group Visits for Chinese Hypertensives Based on Primary Health Care Center |
| *13* | *Gardiner (2017)* | Design of the integrative medical group visits randomized control trial for underserved patients with chronic pain and depression |
| *13* | *Gardiner (2019)* | Effectiveness of integrative medicine group visits in chronic pain and depressive symptoms: A randomized controlled trial |
| *14* | *Liu (2013)* | Effectiveness of using group visit model to support diabetes patient self-management in rural communities of Shanghai: a randomized controlled trial |
| *15* | *Naik (2011)* | Comparative Effectiveness of Goal Setting in Diabetes Mellitus Group Clinics: randomized clinical trial |
| *16* | *Schilinger (2008)* | Seeing in 3-D: Examining the Reach of Diabetes Self-Management Support Strategies in a Public Health Care System |
| *16* | *Schilinger (2009)* | Effects of Self-Management Support on Structure, Process, and Outcomes Among Vulnerable Patients With Diabetes |

| | | |
|---|---|---|
| *16* | *Wallace (2013)* | Influence of Patient Characteristics on Assessment of Diabetes Self-Management Support |
| *17* | *Simon (2015)* | Quality improvement in chronic care delivery for patients with arterial hypertension through Group Medical Visits: Patient acceptance and attendance in the German pilot project |
| *18* | *Taveira (2011)* | Pharmacist-Led Group Medical Appointments for the Management of Type 2 Diabetes with Comorbid Depression in Older Adults |
| *19* | *Taveira (2014)* | Interventions to Maintain Cardiac Risk Control After Discharge from a Cardiovascular Risk Reduction Clinic: A Randomized Controlled Trial |
| *20* | *Vaughan (2017)* | Integrating CHWs as Part of the Team Leading Diabetes Group Visits A Randomized Controlled Feasibility Study |
| *21* | *Vaughan (2020)* | A Telehealth-supported, Integrated care with CHWs, and MEdication-access (TIME) Program for Diabetes Improves HbA1c: a Randomized Clinical Trial |
| *22* | *Wagner (2001)* | Chronic Care Clinics for Diabetes in Primary Care: a system side randomized trial |
| *23* | *Wu (2018)* | Costs and effectiveness of pharmacist-led group medical visits for type-2 diabetes: A multi-center randomized controlled trial |

### S4 – Outcomes reported by studies

Most trials (*n*=19, 83%) included at least one biomedical health indicator, particularly trials of SMAs for a single health condition (see Table S4). The most common measure (*n*=18, 78%), was HbA1C (%) [23,25,26,29–31,33,34,37–44,46,53]. Three of these trials were of SMAs for patients with multiple LTCs – diabetes with CVD risk or diabetes and overweight [44]. Two trials reported the proportion of patients meeting HbA1C goals at 6 months [40,43].

Other biomedical health indicators included systolic and or diastolic blood pressure (DBP) reported in 14 trials [23,26–30,37,38,40,42–49,53]. LDL cholesterol (mg/dL) was measured in 12 trials [23–30,37,38,40,42–49,53]. HDL cholesterol mg/dL was measured in 8 trials [23–29,37,38,41,44–49]. Triglycerides, mg/dL, were measured in 8 trials [23–29,31,37,38,44–49]. BMI, (kg/m$^2$), was measured in six trials[30,31,34,37,38,50]. Change in BMI was reported at 12 months in 2 trials [29,32]. Mean weight loss (Kg) was measured in three trials [31,38,44], percentage of group to achieve weight loss goals (≥5% weight loss/any weight loss/any weight gain) was measured in one trial [37] and change in weight from baseline was reported in another trial [29].

Several trial outcome measures included psychological and well-being measures. Participant self-completion questionnaires were used to measure depression in seven trials [21,22,31,32,39,43,46–50], quality of life in six trials [19–22,31,34–36,42,53], and diabetes self-efficacy in seven trials [19,23,30,33–36,46–50].

There was much heterogeneity in the outcomes reported by studies and measures used to measures these outcomes. See Table A5. Behavioural outcomes included measures of physical activity in six trials [23,31,34,40,44,50], healthcare service use was measured in nine trials [19,22,23,25,29,39,41,43,46] and medication adherence in four trials [40,46,50,52].

Health service costs were measured in seven trials reported as cost per study arm [19,39,52] cost per patient per year [25,26,46], per person [42].

Table S4: All outcomes (and measures) reported by trials

| Trial (condition) | Outcomes (scales/measures) |
|---|---|
| ***Baqir 2020***<br><br>*(Osteoporosis)* | **Biomedical:** NR<br><br>**Psychological:** patient satisfaction with group clinics<br><br>**Behavioural:** MPR (mean possession ratio) with bisphosphonates (MPR is a useful surrogate marker for adherence), persistence with treatment with bisphosphonates in months after treatment was initiated (calculated as the number of months treatment taken before stopping)<br><br>**Costs:** cost differences between the group and 1:1 clinics. |
| ***Berry 2016***<br><br>*(Diabetes)* | **Biomedical:** Blood pressure (systolic and diastolic), A1C %, fasting lipid panel (triglycerides, LDL, HDL), glucose testing, diet, medical care, heart rate (beat/min).<br><br>**Psychological:** Stanford Diabetes Self-Management Questionnaire (general health, symptoms (fatigue, fear, worry, shortness of breath, pain etc.)<br><br>**Behavioural:** Physical activities during the past week, confidence about doing things, daily activities, health care service use<br><br>**Costs:** NR<br><br>**Other**: NR |
| ***Clancy 2003a,b***<br><br>(Diabetes) | **Biomedical**: HbA1c (%), cholesterol (mg/dl), triglycerides (mg/dl), HDL (mg/dl), LDL (mg/dl), Primary Care Assessment Tool [PCAT],<br><br>**Psychological:** Trust in Physician Scale<br><br>**Behaviours:** SMA attendance<br><br>**Cost:** charges (outpatient, inpatient, and emergency room costs and use)<br><br>**Other:** mean total criteria met for 10 process-of-care indicators (up-to-date HbA1c levels and lipid profiles; urine for microalbumin; use of ACE inhibitor or angiotensin receptor blocker, especially in the face of microalbuminuria; use of lipid-lowering agents for LDL levels <100 mg/dl; daily use of aspirin; annual foot examinations; annual referrals for retinal examinations; and immunizations against streptococcal pneumonia and influenza) |
| ***Clancy 2007a,b,2008***<br><br>(Diabetes) | **Biomedical:** HbA1c (%), lipid profiles (total cholesterol, HDL, LDL, triglycerides), blood pressure,<br><br>**Psychological:** Diabetes Locus of Control Survey, trust in the healthcare, provider perception of characteristics<br><br>**Behaviour**: patients' attendance to group visits<br><br>**Costs**: charges (portioned into outpatient visits, emergency department visits, and inpatient stays)<br><br>**Other:** Primary Care Assessment Tool [PCAT] |
| ***Cohen 2011***<br><br>(Diabetes CVD risk) | **Biomedical**: Proportion of patients achieving target goals as recommended by the ADA (A1C, LDL, systolic blood pressure, weight),<br><br>**Psychological**: Quality-of-life questionnaire (VR-36), Perceived Competence<br><br>**Behavioural**: medication adherence (individual medications/any cholesterol medication/total antihypertensive medications/total diabetes medications/total |

| | |
|---|---|
| | cholesterol medications), self care activities (general diet, specific diet, exercise, blood sugar testing, foot care, number of cigarettes smoked per day),<br><br>**Other**: NR |
| ***Cole 2013***<br><br>*(Prediabetes)* | **Biomedical**: Weight, BMI, systolic blood pressure, diastolic blood pressure, HbA1C (%), fasting blood glucose (mg/dl), total cholesterol, LDL, HDL, triglycerides) albumin-creatinine ratio<br><br>**Psychological**: NR<br><br>**Behavioural:** NR<br><br>**Costs:** NR<br><br>**Other**: NR |
| ****Coleman 2001***<br><br>*(Chronic conditions)* | **Biomedical**: NR<br><br>**Psychological**: NR<br><br>**Behavioural:** Percentage of participants who made one or more emergency visits, the average number of emergency visits, repeated emergency department visits, hospitalisations, primary care visits, primary care + group visits, number of group visits attended [intervention participants only]<br><br>**Costs**: NR<br><br>**Other**: NR |
| ****Scott 2004***<br><br>*(Chronic conditions)* | **Biomedical: NR**<br><br>**Psychological and wellbeing**: quality of life, *self-efficacy scale (confidence communicating with my physician, confidence managing my disease, confidence doing chores, confidence with social activities, confidence managing depression)*, patient satisfaction (with PCP, PCP attentiveness, PCP unhurriedness, PCP explanation of condition, time spent with PCP, clinic nurse, overall quality of care, amount of health education), patient satisfaction at 24 months with (talking to PCP about advanced directives, talking with the pharmacist, education with the pharmacist, education from the nurse), functional outcomes (health status and advanced, household, and basic Activities of Daily Living),<br><br>**Behavioural**: Healthcare utilisation (clinic vists/patient; pharmacy fills/patient; hospital admissions/patient; hospital observation admissions/patient; hospital outpatient visits/patient; professional services/patient; emergency visits/patient; skilled nursing facility admissions/patient; home visits/patient), patients' attendance to group visits<br><br>**Costs**: costs of healthcare utilisation (clinic, pharmacy, hospital, hospital observation, hospital outpatient, professional services, emergency room, skilled nursing facility, home health, cost of termination from Kaiser Permanente, total cost) |
| ***Drake 2018***<br><br>(Diabetes) | **Biomedical**: HbA1c, BMI, blood pressure (systolic and diastolic), and low-density lipoprotein<br><br>**Psychological**: Diabetes Empowerment Scale-Short Form (diabetes-specific health self-management skills), General Self-Rated Health (GSRH), PHQ-2 (depression screening tool), visual goal progress scale, patient satisfaction, the 13-item Patient Activation Measure (self-management and patient engagement)<br><br>**Behavioural**: retention<br><br>**Costs**: NR<br><br>**Other**: NR |

| | |
|---|---|
| ****Edelman (2010)**<br><br>*(Diabetes and hypertension)* | **Biomedical**: HbA1c, Diastolic BP, Systolic BP, mean adverse events (hypoglycaemic episodes-falls or light-headedness),<br><br>**Behavioural**: medication adherence, blood pressure control, HbA1C control, Hospital admissions, ED visits (primary care and emergency care visits by using Veterans Affairs–specific codes. Visit counts are exclusive of group clinic sessions), attendance rates<br><br>**Psychological**: perceived competence score: Self-efficacy,<br><br>**Costs**: cost of SMAs<br><br>**Costs**: NR |
| ****Crowley (2013)**<br><br>*(Diabetes and hypertension)* | **Biomedical**: HbA1c (change in HbA1c from study baseline to study end), rates of hypoglycaemia or a self-reported episode of hypoglycaemia<br><br>**Psychological:** Self-efficacy(measured by the Perceived Competence Scale (PCS)),<br><br>**Behavioural:** attendance rates<br><br>**Costs**: NR<br><br>**Other**: NR |
| ****Crowley (2014)**<br><br>*(Diabetes and hypertension)* | **Biomedical**: Mean total cholesterol, LDL-C, high-density lipoprotein cholesterol (HDL-C), and triglyceride levels as continuous outcomes, generated a dichotomous outcome variable indicating whether patients met Adult Treatment Panel III criteria for LDL-C control (LDL-C 400 mg/dL, LDL-C was not reported.<br><br>**Psychological**: NR<br><br>**Behavioural**: medication use (intensification of LDL-C-lowering medications during the study period)<br><br>**Costs**: NR<br><br>**Other**: NR |

| | |
|---|---|
| ***Ee 2020***<br><br>*(Diabetes)* | **Biomedical:** HbA1c (also time in range/mean blood glucose); Anthropometric measures (weight, BMI, and hip circumference, and waist/hip circumference ratio), Blood pressure (systolic and diastolic), Total cholesterol, Triglycerides. Glycemic management, Fasting lipids<br><br>**Psychological:** depression (Becks depression inventory), Quality of Life (EQ5D5L sub groups: anxiety, mobility, pain discomfort, self-care, usual activity, Visual acuity scale), Anxiety (State Anxiety Index) Diabetes-related distress, Patient-Reported Outcomes Information System questionnaire (PROMIS-29)<br><br>**Behavioural:** Self-reported diet quality (daily serves of vegetables and fruit, weekly serves of takeaways) from the Population Health Survey Questionnaire, Physical activity levels, Number of minutes of mindfulness practice per week, Number of minutes of mindfulness practice per week, Recruitment rates, Retention rate<br><br>**Costs:** NR<br><br>**Other:** Patient-Reported Outcomes Information System questionnaire (PROMIS-29) (Pain intensity, anxiety, depression, fatigue, pain interference, physical function, sleep, social) , |
| ***Gao 2015***<br><br>*(Hypertension)* | **Biomedical:** Systolic BP, Diastolic BP, BMI,<br><br>**Psychological and wellbeing:** Self-reported health, energy, Self-efficacy of managing symptoms/disease/ physical activity, attitudes, depression, health distress<br><br>**Behavioural:** treatment compliance (never taking medicine, taking without compliance, taking with compliance), physical activities, dietary adjusting (NA, never adjusting; AWOC, adjusting without compliance; AWC, adjusting with compliance.)<br><br>**Costs**: NR<br><br>**Other:** patient-physician communication, social support, coping skills, beliefs |
| ***Gardiner 2019***<br><br>*(Chronic pain and depression)* | **Biomedical**: Brief Pain Inventory (Assesses pain severity and interference, average pain score in last 7 days), Pain,<br><br>**Psychological and wellbeing**: Patient Health Questionnaire (PHQ-9) a self-reported depression scale, Pain Self Efficacy Questionnaire (PSEQ) Patient Activation Measure, Short Form 12 Health Survey QoL, Common opioid misuse measure, Perceived Stress Scale.<br><br>**Behavioural**: Self- Reported Pain Medication Use (past 7 days), ED use<br><br>**Costs**: NR<br><br>**Other**: NR |
| ***Liu (2013)***<br><br>*(Diabetes)* | **Biomedical:** BMI, systolic BP, diastolic BP<br><br>**Psychological and wellbeing:** Self-management behaviour scores (diet, aerobic exercise, practice of cognitive symptom management, communication with doctor, examining feet), Diabetes Self-Efficacy Scale, health status (self-rated health, energy, health distress, fatigue, illness intrusiveness, depression)<br><br>**Behavioural:** NR<br><br>**Costs**: NR<br><br>**Other**: NR |

| | |
|---|---|
| ***Naik (2011)***<br>*(Diabetes)* | **Biomedical:** HbA1C (%), [BMI and systolic BP only measured at baseline]<br><br>**Psychological:** Diabetes Self-Efficacy<br><br>**Behavioural: NR**<br><br>**Costs:** NR<br><br>**Other**: NR |
| *******Schilinger (2008/09)***<br><br>*(Diabetes)* | **Biomedical:** HbA1C (%), BMI, blood pressure, BMI<br><br>**Psychological and wellbeing**: Diabetes Quality Improvement Programe Diabetes Self-Efficacy, weekly self-care, quality of life (Short Form-12) functional status (restricted activity; bed days, prior month),<br><br>**Costs**: NR<br>**Behavioural**: exercise (estimated minutes of moderate and vigorous physical activity on each of the days)<br><br>**Other**: reach (participant among clinics, clinicians, and patients; patient representativeness; patient engagement with SMA), structure of care, Patient Assessment of Chronic Illness Care, patient reports of providers' communication over the prior year (Interpersonal Processes of Care for Diverse Populations), |
| ******Wallace 2013***<br><br>*(Diabetes)* | **Biomedical**: HbA1C, Systolic BP, Diastolic BP, BMI, Behavioural and Functional outcomes (spent most of the day in bed due to health problems" and the extent to which diabetes prevented them from carrying out normal daily activities (diabetes interference),<br><br>**Psychological wellbeing:** quality of life (Short Form (SF)-12 instrument), Test of Functional Health Literacy in Adults (0-36), Social support network, Health Status<br><br>**Behavioural**: self-management behaviours (eating healthy foods, following a diabetic diet, exercising, self-monitoring of blood glucose, and caring for one's feet)<br><br>**Other: PACIC rating**. (The PACIC is a questionnaire consisting of 26 items with responses ranging from almost never (0) to almost always (5), includes: Delivery System Design/ Decision Support (e.g., Given a written list of things I should do to improve my health), Goal Setting (e.g., Asked to talk about my goals in caring for my illness), Problem-Solving/ Contextual Counseling (e.g., Helped make a treatment plan that I could do in my daily life), and Follow-Up/Coordination (e.g., Contacted after a visit to see how things were going). Subscale scores are calculated as means of the items within each subscale. Patient Activation<br><br>**Costs:** NR |
| ***Simon & Sawicki 2015***<br><br>*(Hypertension)* | **Biomedical:** NR<br><br>**Psychological and wellbeing**: Patients' willingness to attend SMA<br><br>**Behavioural**: SMA attendance<br><br>**Costs**: NR<br><br>**Other**: NR |

| | |
|---|---|
| ***Taveira (2011)***<br><br>*(Diabetes and depression)* | **Biomedical:** Attained a goal of A1C of 7%, proportion of participants who attained the ADA guideline recommendations for blood pressure and fasting lipid levels, 10-year risk of cardiac events,<br><br>**Psychological and wellbeing**: Perceived Competence Diabetes Scale, Patient Health Questionnaire-9 Depression Scale<br><br>**Behavioural:** Summary of Diabetes Self-Care Activities, number of patients who used tobacco (average daily number of cigarettes), medication changes (dose increase or initiation of any antihypertensive/antihyperglycemic agent/antihyperlipidemic/antidepressants), healthcare utilisation (primary care provider visits, emergency department visits, hospital admission rates),<br><br>**Costs:** NR<br><br>**Other:** deaths |
| ***Taveira 2014***<br><br>*(Diabetes and CVD risk)* | **Biomedical**: Time to failure for guideline recommended goals of HbA1c and BP, Maintenance of goals for LDL, change in prescribed medications<br><br>**Psychological and wellbeing**: NR<br><br>**Behavioural:** hospital admissions and ED visits<br><br>**Costs**: NR<br><br>**Other**: NR |
| ***Vaughan 2017***<br><br>*(Diabetes)* | **Biomedical**: HbA1C (not measured for participants with pre-diabetes, achieved target A1C%)**,** weight (> 5% weight loss), lipid levels (total cholesterol, HDL, LDL, triglyerides, SBP, DBP, BMI<br><br>**Psychological and wellbeing:** NR<br><br>**Behavioural:** NR<br><br>**Costs:** NR<br><br>**Other**: 8 standards of care per ADA and USPTF: weight loss, lipid levels, retinal screening, foot exam (i.e., assessment of foot pulses, sensation, skin exam), urine microalbumin, cancer screening (breast, cervical, colorectal) |
| ***Wagner 2001***<br>*(Diabetes)* | **Biomedical**: HbA1C (%), cholesterol (mean mg/dl)<br><br>**Psychological and wellbeing**: health status (general health, physical function, physical role limitation, bed disability days, restricted activity day, depression (Center for Epidemiological Studies Depression Scale), satisfaction (medical care satisfaction, diabetes care satisfaction)<br><br>**Behavioural**: healthcare utilisation (primary care visits, ER visits, speciality visits, hospital admissions)<br><br>**Cost:** NR<br><br>**Other**: receipt of recommended preventative manoeuvres (prevention procedures, medication review, retinal eye exam, foot examination, microalbumin test), use and helpfulness of patient education (written material, classes, face-to-face counselling) |

| | |
|---|---|
| ***Wu 2018***<br><br>*(Diabetes and CVD)* | **Biomedical**: Coronary Event Risk score, HbA1c %, systolic BP, LDL,<br><br>**Psychological and wellbeing**: health-related quality-of-life (veteran version of Medical Outcomes Study survey (SF-36v)<br><br>**Behavioural:** NR<br><br>**Costs**: healthcare costs (provider time in the group medical visits, medications, hospitalisations, ED visits, lab tests, procedures, referrals, outpatient clinic visits) |
| ***Yancy 2020***<br><br>*(Diabetes and overweight)* | **Biomedical**: HbA1c, hypoglycemic events, diabetes medication use, Weight, SBP, DBP, total cholesterol, LDL, HDL, waist circumference, triglycerides.<br><br>**Psychological and wellbeing**: diabetes related emotional distress (Problem Areas in Diabetes-PAID)<br><br>**Behavioural**: Dietary adherence, physical activity, medication nonadherence<br><br>**Cost**: Cost-effectiveness |

*Same trial

### S5 – Quality assessment of included studies with sensitivity analyses

Table S5a: Quality assessments conducted for each included trial

| Study | Domain | | | | | | |
|---|---|---|---|---|---|---|---|
| | Selection bias | | Performance bias | Detection bias | Attrition bias | Reporting bias | Other bias |
| | ***Random sequence generation*** | ***Allocation concealment*** | ***Blinding of participants and personnel*** | ***Blinding of outcome assessment*** | ***Incomplete outcome data*** | ***Selective reporting*** | ***Other sources of bias*** |
| *Baqir (2016)* | Low | Low | High | Low | Low | Unclear | Low |
| *Berry (2016)* | Unclear | Unclear | Unclear | Unclear | Unclear | Unclear | Unclear |
| Clancy (2003) | Low | Low | High | Low | Low | Unclear | Low |
| Clancy (2007) | Low | Low | High | Unclear | Low | Unclear | Low |
| Cohen (2011) | Unclear | Unclear | High | Unclear | Unclear | High | Low |
| Cole (2013) | Low | Low | High | Unclear | Low | Unclear | Low |
| *Drake (2018)* | Low | Unclear | High | Unclear | High | Low | High |
| Edelman (2010) | Unclear | Unclear | High | Low | Low | Low | Low |
| *Ee (2020)* | Low | Low | High | Low | Low | Low | Low |
| *Gao (2015)* | Unclear | Unclear | Unclear | Low | High | Unclear | High |
| *Gardiner (2019)* | Low | Low | High | Low | Low | Low | Low |
| Liu (2012) | Low | Unclear | High | Low | Unclear | Low | Low |
| Naik (2011) | Unclear | Low | High | Unclear | Unclear | Low | Low |

| | | | | | | | |
|---|---|---|---|---|---|---|---|
| Schillinger (2008) | Unclear | Unclear | High | Unclear | Low | Unclear | Low |
| Scott (2004) | Low | Unclear | High | Unclear | Low | Unclear | High |
| Simon (2015) | Unclear | Unclear | Unclear | Unclear | Unclear | Unclear | Unclear |
| Taveria (2011) | Low | Unclear | High | Unclear | Low | Unclear | Low |
| *Taveira (2014)* | Unclear | Unclear | High | Unclear | High | Low | Low |
| *Vaughan (2017)* | Unclear | Unclear | Unclear | Unclear | Low | Unclear | Low |
| Vaughan (2020) | Low | Unclear | High | Unclear | Unclear | Low | Low |
| Wagner (2001) | Unclear | Unclear | High | Unclear | Unclear | Unclear | Unclear |
| *Wu (2018)* | Low | Low | High | Low | High | High | Low |
| *Yancy (2020)* | Low | Low | High | Low | High | Low | High |

Figure S5: Summary of each of risk bias

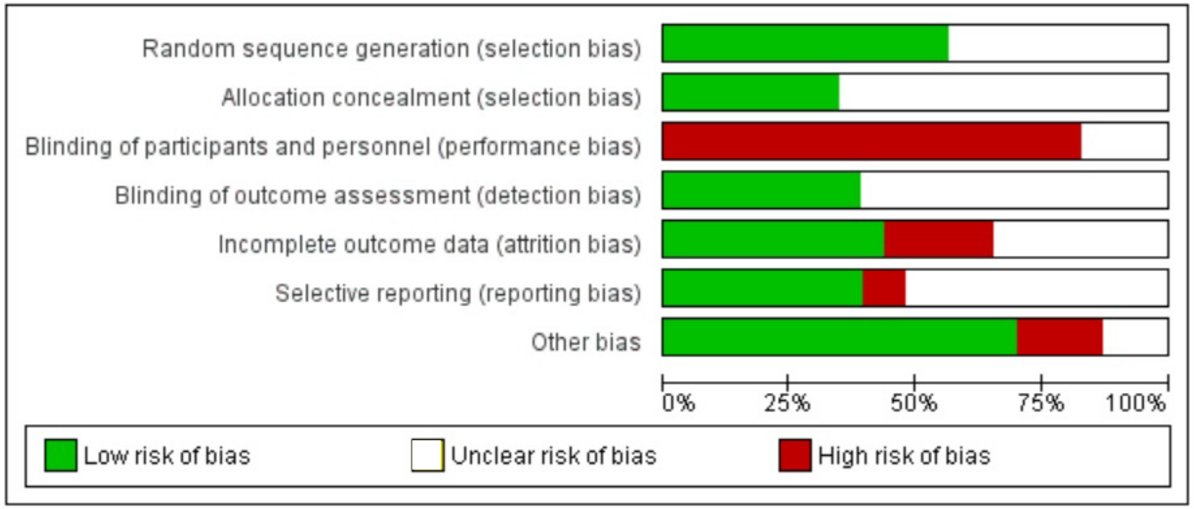

**Sensitivity Analyses according to Risk of Bias Criteria**

Table S5a: Comparison between mean effect sizes for HbA1c (%) according to risk of bias across domains

| Domain | Post-intervention changes in HbA1c (%) (*k*=8) | | | | | |
|---|---|---|---|---|---|---|
| | Low risk | | High/Unclear risk | | Confidence Intervals | Sig |
| | *n* | *d* | *n* | *d* | | |
| **Selection bias** | | | | | | |
| Random sequence generation | 6 | -0.074 | 3 | -0.147 | -0.74, -0.59 | 0.798 |
| Allocation concealment | 4 | 0.140 | 5 | -0.263 | -0.93, 0.14 | 0.126 |
| **Performance bias** | | | | | | |
| Blinding of participants and personnel | N/A – all high/unclear | | | | | |
| **Detection bias** | | | | | | |
| Blinding of outcome assessment | 2 | 0.088 | 7 | -0.158 | -1.09, 0.49 | 0.404 |
| **Attrition bias** | | | | | | |
| Incomplete outcome data | 5 | -0.036 | 4 | -0.169 | -0.77, 0.49 | 0.620 |
| **Reporting bias** | | | | | | |
| Selective reporting | 3 | -0.027 | 6 | -0.136 | -0.83, 0.58 | 0.691 |

*n*=frequencies, *k*=number of tests of relationships, *d*=mean effect size
*$p < 0.05$; **$p < 0.01$; ***$p < 0.001$.

Table S5b: Comparison between mean effect sizes for diastolic blood pressure according to risk of bias across domains

| Domain | Post-intervention changes in diastolic blood pressure (*k*=8) | | | | | |
|---|---|---|---|---|---|---|
| | Low risk | | High/Unclear risk | | Confidence Intervals | Sig |
| | *n* | *d* | *n* | *d* | | |
| **Selection bias** | | | | | | |
| Random sequence generation | 3 | -0.201 | 5 | -0.115 | -0.31, 0.48 | 0.617 |
| Allocation concealment | 2 | -0.052 | 6 | -0.126 | -0.63, 0.49 | 0.771 |
| **Performance bias** | | | | | | |
| Blinding of participants and personnel | N/A – all high/unclear | | | | | |
| **Detection bias** | | | | | | |
| Blinding of outcome assessment | 2 | -0.080 | 6 | -0.210 | -0.37, 0.11 | 0.241 |
| **Attrition bias** | | | | | | |
| Incomplete outcome data | 4 | -0.189 | 4 | -0.109 | -0.21, 0.37 | 0.528 |
| **Reporting bias** | | | | | | |
| Selective reporting | 3 | -0.089 | 5 | -0.127 | -0.41, 0.32 | 0.766 |

*n*=frequencies, *k*=number of tests of relationships, *d*=mean effect size
**p*< 0.05; ***p*< 0.01; ****p*< 0.001.

Table S5c: Comparison between mean effect sizes for total cholesterol according to risk of bias across domains

| Domain | Post-intervention changes in total cholesterol (*k*=3) | | | | | |
|---|---|---|---|---|---|---|
| | Low risk | | High/Unclear risk | | Confidence Intervals | Sig |
| | *n* | *d* | *n* | *d* | | |
| **Selection bias** | | | | | | |
| Random sequence generation | 2 | 0.014 | 1 | 0.021 | -4.02, 4.04 | 0.985 |
| Allocation concealment | 1 | -0.422 | 2 | 0.032 | -5.86, 6.77 | 0.529 |
| **Performance bias** | | | | | | |
| Blinding of participants and personnel | N/A – all high/unclear | | | | | |
| **Detection bias** | | | | | | |
| Blinding of outcome assessment | 1 | -0.422 | 2 | 0.032 | -5.86, 6.77 | 0.529 |
| **Attrition bias** | | | | | | |
| Incomplete outcome data | 1 | -0.422 | 2 | 0.032 | -5.86, 6.77 | 0.529 |
| **Reporting bias** | | | | | | |
| Selective reporting | 2 | 0.014 | 1 | 0.021 | -4.02, 4.04 | 0.985 |

Table S5d: Comparison between mean effect sizes for high-density lipoprotein cholesterol according to risk of bias across domains

| Domain | Post-intervention changes in high-density lipoprotein cholesterol (*k*=3) | | | | | |
|---|---|---|---|---|---|---|
| | Low risk | | High/Unclear risk | | Confidence Intervals | Sig |
| | *n* | *d* | *n* | *d* | | |
| **Selection bias** | | | | | | |
| Random sequence generation | 1 | -0.050 | 2 | 0.504 | -3.33, 4.44 | 0.321 |
| Allocation concealment | 1 | -0.050 | 2 | 0.504 | -3.33, 4.44 | 0.321 |
| **Performance bias** | | | | | | |
| Blinding of participants and personnel | N/A – all high/unclear | | | | | |
| **Detection bias** | | | | | | |
| Blinding of outcome assessment | N/A – all high/unclear | | | | | |
| **Attrition bias** | | | | | | |
| Incomplete outcome data | 2 | 0.170 | 1 | 0.550 | -4.59, 5.35 | 0.509 |
| **Reporting bias** | | | | | | |
| Selective reporting | N/A – all high/unclear | | | | | |

*n*=frequencies, *k*=number of tests of relationships, *d*=mean effect size
*$p$< 0.05; **$p$< 0.01; ***$p$< 0.001.

Table S5e: Comparison between mean effect sizes for low-density lipoprotein cholesterol according to risk of bias across domains

| Domain | Post-intervention changes in low-density lipoprotein cholesterol (*k*=5) | | | | | |
|---|---|---|---|---|---|---|
| | Low risk | | High/Unclear risk | | Confidence Intervals | Sig |
| | *n* | *d* | *n* | *d* | | |
| **Selection bias** | | | | | | |
| Random sequence generation | 2 | 0.042 | 3 | -0.061 | -1.03, 0.88 | 0.812 |
| Allocation concealment | 2 | 0.042 | 3 | -0.061 | -1.03, 0.88 | 0.812 |
| **Performance bias** | | | | | | |
| Blinding of participants and personnel | N/A – all high/unclear | | | | | |
| **Detection bias** | | | | | | |
| Blinding of outcome assessment | 1 | 0.140 | 4 | -0.097 | -1.18, 0.71 | 0.482 |
| **Attrition bias** | | | | | | |
| Incomplete outcome data | 2 | 0.067 | 3 | -0.072 | -1.14, 0.86 | 0.685 |
| **Reporting bias** | | | | | | |
| Selective reporting | 1 | -0.020 | 4 | -0.034 | -1.18, 1.15 | 0.971 |

*n*=frequencies, *k*=number of tests of relationships, *d*=mean effect size
*$p < 0.05$; **$p < 0.01$; ***$p < 0.001$.

Table S5f: Comparison between mean effect sizes for triglycerides according to risk of bias across domains

| **Domain** | **Post-intervention changes in triglycerides (*k*=4)** | | | | | |
|---|---|---|---|---|---|---|
| | **Low risk** | | **High/Unclear risk** | | **Confidence Intervals** | **Sig** |
| | ***n*** | ***d*** | ***n*** | ***d*** | | |
| **Selection bias** | | | | | | |
| Random sequence generation | 2 | -0.210 | 1 | -1.111 | -5.27, 3.47 | 0.232 |
| Allocation concealment | 1 | 0.118 | 2 | -0.732 | -10.96, 9.26 | 0.479 |
| **Performance bias** | | | | | | |
| Blinding of participants and personnel | N/A – all high/unclear | | | | | |
| **Detection bias** | | | | | | |
| Blinding of outcome assessment | 1 | 0.118 | 2 | -0.732 | -10.96, 9.26 | 0.479 |
| **Attrition bias** | | | | | | |
| Incomplete outcome data | 1 | 0.118 | 2 | -0.732 | -10.96, 9.26 | 0.479 |
| **Reporting bias** | | | | | | |
| Selective reporting | 2 | -0.210 | 1 | -1.111 | -5.27, 3.47 | 0.232 |

*n*=frequencies, *k*=number of tests of relationships, *d*=mean effect size
**p*< 0.05; ***p*< 0.01; ****p*< 0.001.

Table S5g: Comparison between mean effect sizes for BMI according to risk of bias across domains

| Domain | Post-intervention changes in BMI (*k*=5) | | | | | |
|---|---|---|---|---|---|---|
| | Low risk | | High/Unclear risk | | Confidence Intervals | Sig |
| | *n* | *d* | *n* | *d* | | |
| **Selection bias** | | | | | | |
| Random sequence generation | 2 | 0.278 | 3 | 0.039 | -0.-90, 1.09 | 0.789 |
| Allocation concealment | 1 | 0.955 | 4 | 0.007 | -2.84, 1.01 | 0.226 |
| **Performance bias** | | | | | | |
| Blinding of participants and personnel | | | N/A – all high/unclear | | | |
| **Detection bias** | | | | | | |
| Blinding of outcome assessment | 2 | 0.361 | 3 | -0.095 | -1.32, 0.69 | 0.386 |
| **Attrition bias** | | | | | | |
| Incomplete outcome data | 3 | 0.101 | 2 | 0.000 | -1.18, 0.97 | 0.771 |
| **Reporting bias** | | | | | | |
| Selective reporting | 2 | 0.278 | 3 | 0.039 | -0.90, 1.09 | 0.789 |

*n*=frequencies, *k*=number of tests of relationships, *d*=mean effect size
*$p < 0.05$; **$p < 0.01$; ***$p < 0.001$.

Table S5h: Comparison between mean effect sizes for self-efficacy according to risk of bias across domains

| Domain | Post-intervention changes in self-efficacy (*k*=4) | | | | | |
|---|---|---|---|---|---|---|
| | Low risk | | High/Unclear risk | | Confidence Intervals | Sig |
| | *n* | *d* | *n* | *d* | | |
| **Selection bias** | | | | | | |
| Random sequence generation | 1 | -0.282 | 3 | 0.373 | -0.11, 1.42 | 0.066 |
| Allocation concealment | 1 | -0.282 | 3 | 0.373 | -0.11, 1.42 | 0.066 |
| **Performance bias** | | | | | | |
| Blinding of participants and personnel | N/A – all high/unclear | | | | | |
| **Detection bias** | | | | | | |
| Blinding of outcome assessment | 2 | 0.069 | 3 | 0.336 | -1.27, 1.78 | 0.547 |
| **Attrition bias** | | | | | | |
| Incomplete outcome data | 2 | 0.033 | 2 | 0.380 | -0.99, 1.65 | 0.396 |
| **Reporting bias** | | | | | | |
| Selective reporting | 1 | -0.282 | 3 | 0.373 | -0.11, 1.42 | 0.066 |

*n*=frequencies, *k*=number of tests of relationships, *d*=mean effect size
*$p$< 0.05; **$p$< 0.01; ***$p$< 0.001.

Table S5i: Comparison between mean effect sizes for depression according to risk of bias across domains

| **Domain** | **Post-intervention changes in depression (*k*=4)** | | | | | |
|---|---|---|---|---|---|---|
| | **Low risk** | | **High/Unclear risk** | | **Confidence Intervals** | **Sig** |
| | ***n*** | ***d*** | ***n*** | ***d*** | | |
| **Selection bias** | | | | | | |
| Random sequence generation | 2 | -0.326 | 2 | -0.136 | -1.04, 1.35 | 0.628 |
| Allocation concealment | 2 | -0.326 | 2 | -0.136 | -1.04, 1.35 | 0.628 |
| **Performance bias** | | | | | | |
| Blinding of participants and personnel | N/A – all high/unclear | | | | | |
| **Detection bias** | | | | | | |
| Blinding of outcome assessment | 3 | -0.265 | 1 | 0.006 | -1.34, 0.68 | 0.103 |
| **Attrition bias** | | | | | | |
| Incomplete outcome data | 2 | -0.326 | 2 | -0.136 | -1.04, 1.35 | 0.628 |
| **Reporting bias** | | | | | | |
| Selective reporting | 2 | -0.326 | 2 | -0.136 | -1.04, 1.35 | 0.628 |

*n*=frequencies, *k*=number of tests of relationships, *d*=mean effect size
*$p$< 0.05; **$p$< 0.01; ***$p$< 0.001.

Table S5j: Comparison between mean effect sizes for hospital admissions according to risk of bias across domains

| Domain | Post-intervention changes in hospital admissions (*k*=3) | | | | | |
|---|---|---|---|---|---|---|
| | Low risk | | High/Unclear risk | | Confidence Intervals | Sig |
| | *n* | *d* | *n* | *d* | | |
| **Selection bias** | | | | | | |
| Random sequence generation | 1 | -0.277 | 2 | 0.173 | -1.95, 2.85 | 0.253 |
| Allocation concealment | N/A – all high/unclear | | | | | |
| **Performance bias** | | | | | | |
| Blinding of participants and personnel | N/A – all high/unclear | | | | | |
| **Detection bias** | | | | | | |
| Blinding of outcome assessment | N/A – all high/unclear | | | | | |
| **Attrition bias** | | | | | | |
| Incomplete outcome data | 1 | -0.277 | 2 | 0.173 | -1.95, 2.85 | 0.253 |
| **Reporting bias** | | | | | | |
| Selective reporting | 1 | 0.294 | 2 | -0.204 | -3.51, 2.51 | 0.283 |

*n*=frequencies, *k*=number of tests of relationships, *d*=mean effect size
**p*< 0.05; ***p*< 0.01; ****p*< 0.001.

Table S5k: Comparison between mean effect sizes for emergency department use according to risk of bias across domains

| Domain | Post-intervention changes in emergency department use (*k*=4) | | | | | |
|---|---|---|---|---|---|---|
| | Low risk | | High/Unclear risk | | Confidence Intervals | Sig |
| | *n* | *d* | *n* | *d* | | |
| **Selection bias** | | | | | | |
| Random sequence generation | 1 | -0.310 | 3 | 0.048 | -0.23, 0.94 | 0.118 |
| Allocation concealment | N/A – all high/unclear | | | | | |
| **Performance bias** | | | | | | |
| Blinding of participants and personnel | N/A – all high/unclear | | | | | |
| **Detection bias** | | | | | | |
| Blinding of outcome assessment | N/A – all high/unclear | | | | | |
| **Attrition bias** | | | | | | |
| Incomplete outcome data | 1 | -0.310 | 3 | 0.048 | -0.23, 0.94 | 0.118 |
| **Reporting bias** | | | | | | |
| Selective reporting | 1 | 0.000 | 3 | -0.109 | -1.40, 1.18 | 0.756 |

*n*=frequencies, *k*=number of tests of relationships, *d*=mean effect size
**p*< 0.05; ***p*< 0.01; ****p*< 0.001.

Table S5l: Comparison between mean effect sizes for primary care visits according to risk of bias across domains

| Domain | Post-intervention changes in primary care visits (*k*=3) | | | | | |
|---|---|---|---|---|---|---|
| | Low risk | | High/Unclear risk | | Confidence Intervals | Sig |
| | *n* | *d* | *n* | *d* | | |
| **Selection bias** | | | | | | |
| Random sequence generation | 2 | -0.035 | 1 | 0.074 | -1.52, 1.74 | 0.552 |
| Allocation concealment | N/A – all high/unclear | | | | | |
| **Performance bias** | | | | | | |
| Blinding of participants and personnel | N/A – all high/unclear | | | | | |
| **Detection bias** | | | | | | |
| Blinding of outcome assessment | N/A – all high/unclear | | | | | |
| **Attrition bias** | | | | | | |
| Incomplete outcome data | 2 | -0.035 | 1 | 0.074 | -1.52, 1.74 | 0.552 |
| **Reporting bias** | | | | | | |
| Selective reporting | N/A – all high/unclear | | | | | |

*n*=frequencies, *k*=number of tests of relationships, *d*=mean effect size
*$p$< 0.05; **$p$< 0.01; ***$p$< 0.001.

### S6 – Additional biomedical measures examined with forest plots

*Total cholesterol*

Five studies trials reported total cholesterol levels: four reported milligrams per deciliter (mg/dL) [25,37,39,46], and one reported millimoles per liter (mmol/L) [31]. Three of these trials which compared SMA to usual care were pooled together into a meta-analysis [31,37,39] where no statistically significant effect was found (*d*=0.022, *95%CI* = -0.12, 0.17, *k*=3, p=.767) (Figure S6a). Of the two studies which could not be included into the meta-analysis [25,46], only Edelman et al. (2010) reported significant group differences, whereby participants in the SMA group demonstrated lower total cholesterol levels at follow-up (153.9 mg/dL) than those in usual care (168.1 mg/dL) [46].

*High density lipoprotein cholesterol*

Five trials reported high-density lipoprotein (HDL) cholesterol [23,25,29,37,46]. Three of these studies were pooled into a meta-analysis [23,29,37]. A non-statistically significant small difference was found for HDL cholesterol between SMA and usual care at follow-up (*d*=0.312, *95%CI* = -0.06, 0.68, *k*=3, p=0.100) (Figure S6b). Moderate levels of heterogeneity were found ($I^2$=41.0%). Of the other trials which could not be included into the meta-analysis [25,46], Edelman et al. (2010) reported that participants in the SMA group demonstrated statistically significantly lower HDL at follow-up (39.3 mg/dL) than those in usual care (41.3 mg/dL) [46].

*Low-density lipoprotein cholesterol*

Nine trials reported low-density lipoprotein (LDL) cholesterol [23,25,26,29,37,40–42,46]. It was feasible to pool five of these studies [23,29,37,41,42]. SMAs showed no statistically significant difference to usual care on LDL cholesterol at follow-up (*d*=-0.027, *95%CI* = -0.27, 0.22, *k*=5, *p*=0.830) (Figure S6c). Heterogeneity was moderate with an overall $I^2$ of 44.7%. Of the trials which could not be included into the meta-analysis, only Edelman et al. (2010) which looked at SMAs for patients with diabetes and hypertension, found that LDL was significantly lower for participants in the SMA group (*M*=84.1) than the control group (*M*=93.3) at follow-up [46].

*Triglycerides*

Triglycerides were measured in seven trials: mmol/L [31], mg/dL [23,25,26,29,37] and log mg/dL [46]. It was possible to meta-analyse three of these trials [23,31,37] which compared SMA to usual care. A small but non-statistically significant effect was found (*d*=-0.517, *95%CI* = -1.22, 0.19, *k*=3, p=0.149) (Figure S6d). Of the other four trials which could not be included in the meta-analysis, none reported any between-group differences for triglycerides at follow-up.

*Weight*

Weight (mean kg) was measured in five trials: [29,31,38,40,44]. A small but non-statistically significant effect was found (*d*=0.298, *95%CI* = -0.89, 1.49, *k*=2) (p=0.623) (Figure S6e). High levels of heterogeneity were observed ($I^2$=79.3%). No significant group differences in weight were reported by the three other studies which could not be included into the meta-analysis [29,40,44].

*BMI*

Seven trials measured BMI kg/m$^2$ [29,31,32,34,37,38,50]. There was sufficient data to include four trials on diabetes [31,34,37,38] and a trial of patients with hypertension [50] in a meta-analysis. No statistically significant difference between SMAs and usual care was found for BMI (*d*=0.016, *95%CI* = -0.19, 0.22, *k*=5) (*p*=0.876) (Figure S6f). The pooled effect remained similar when Ee et al. (2020) [31] as removed as an outlier in the sensitivity analysis due to wide 95% CIs, (*d*=-0.007, *95%CI* = -0.15, 0.16, k=4)

None of the two trials which could not be included into the meta-analysis reported any significant differences between the SMA and control groups for BMI at follow-up [29,32].

Figure S6a: Forest plot for total cholesterol

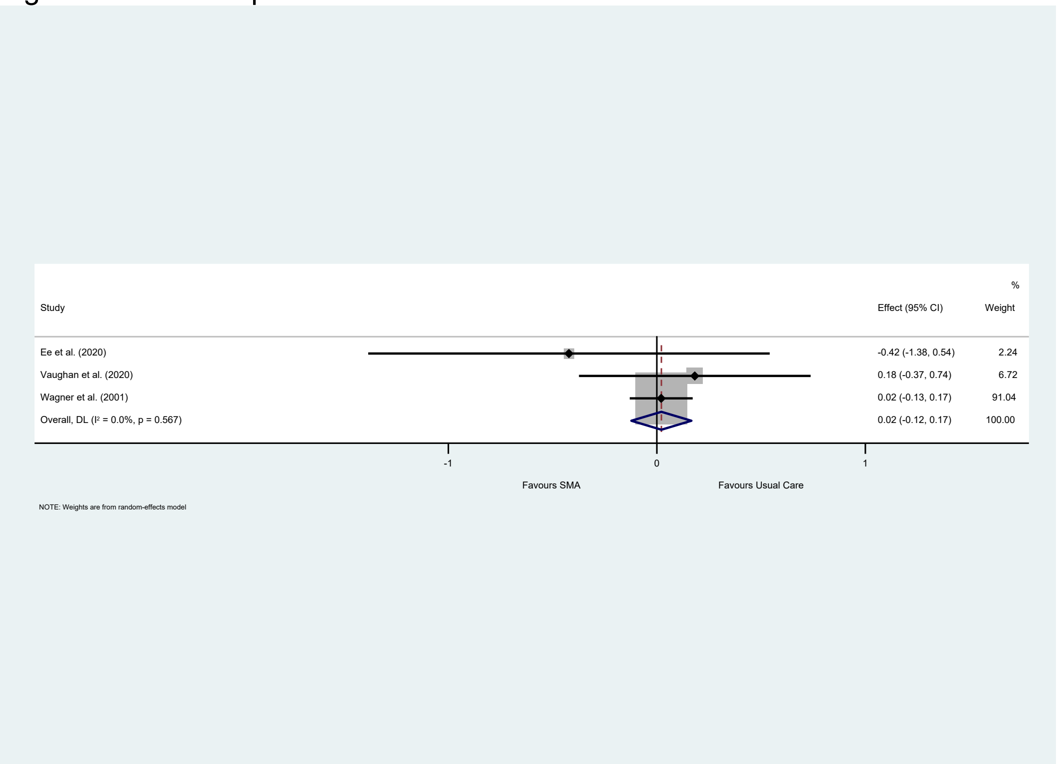

Figure S6b: Forest plot for HDL

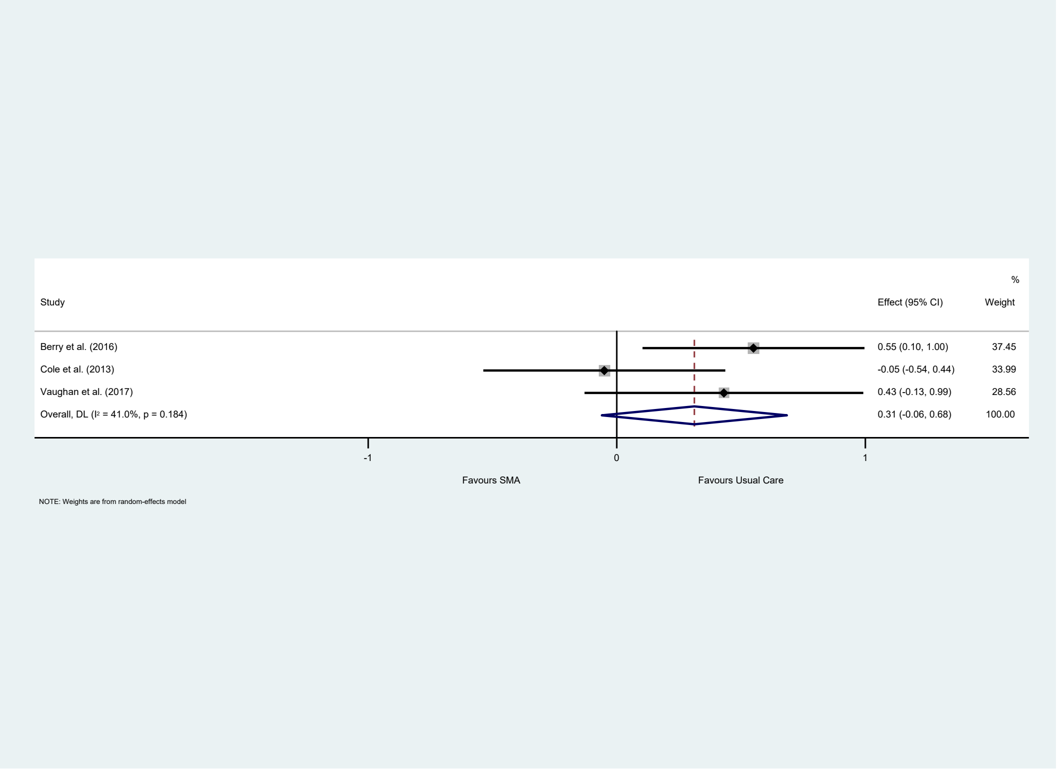


Figure S6c: Forest plot for LDL cholesterol

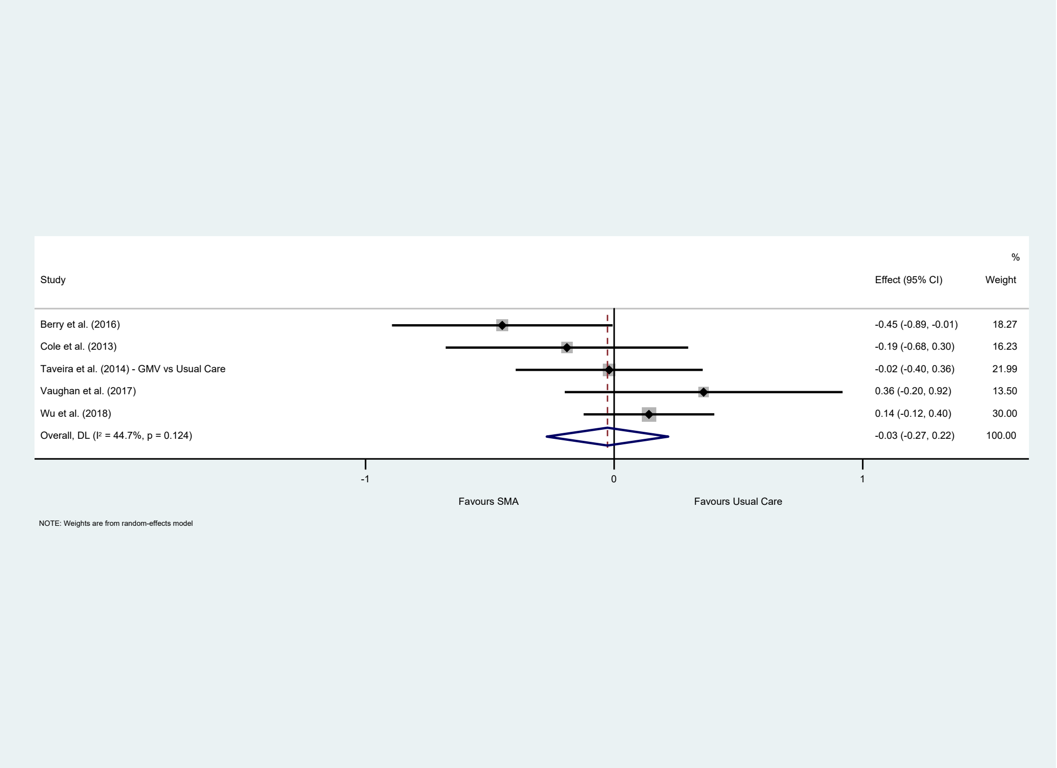

Figure S6d: Forest plot for triglycerides

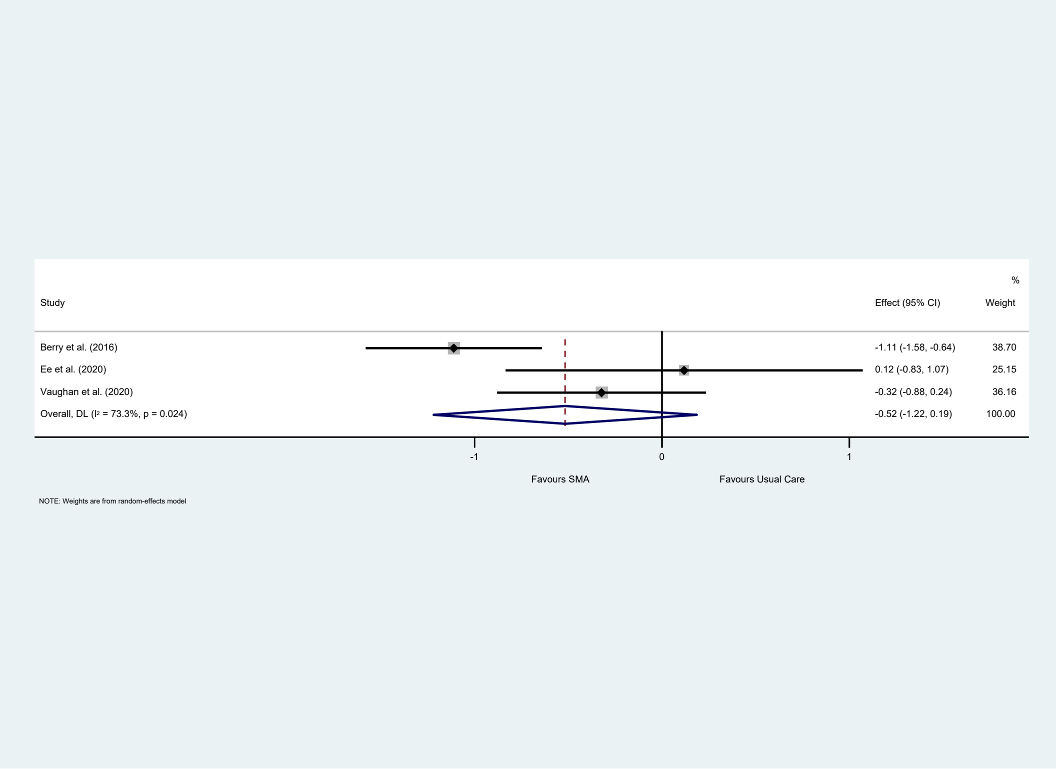


Figure S6e: Forest plot for weight

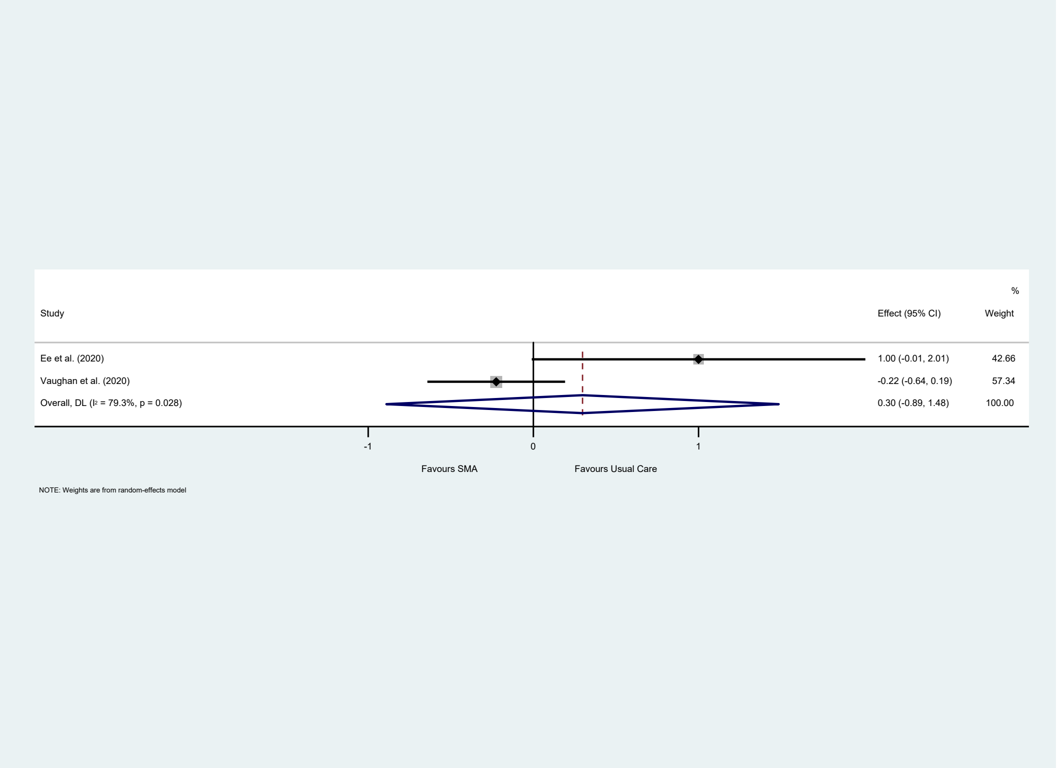

Figure S6f: Forest plot for BMI

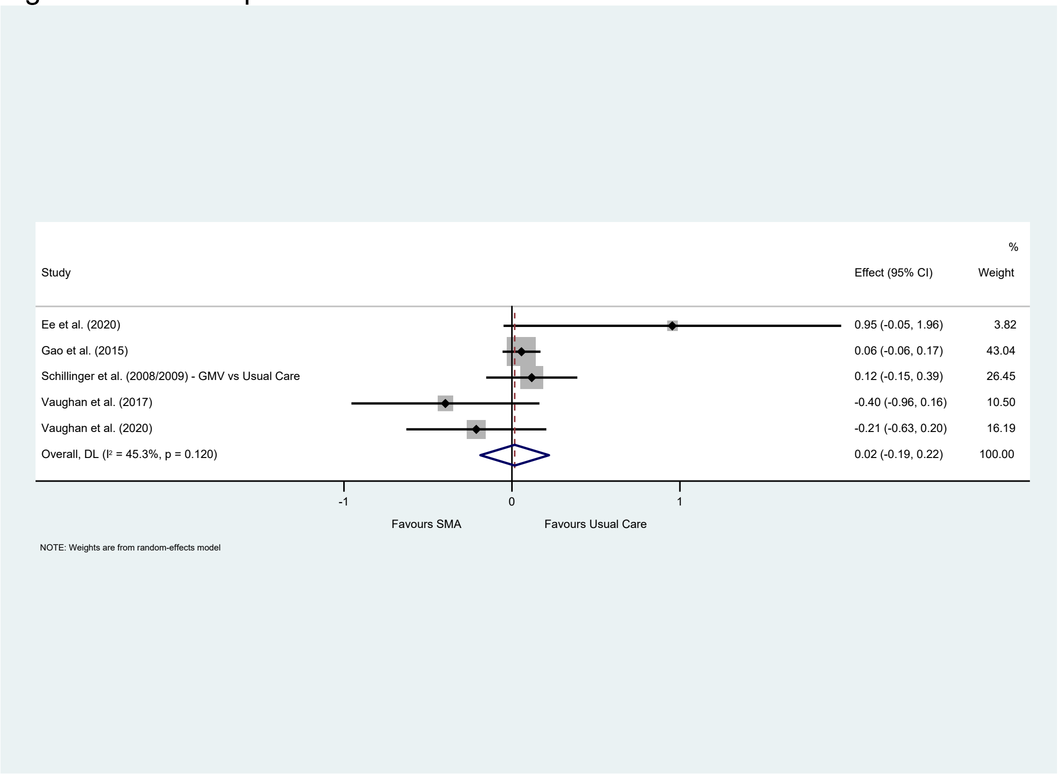

### S7 – Additional psychological and well-being measures examined with forest plots

*Depression*

Of the six trials which reported on depression [22,31,32,39,43,50], it was possible to include four in a meta-analysis [22,31,39,50]. No statistically significance difference between SMAs and usual care for depression was found at follow-up ($d$=-0.173, *95%CI* = -0.38, 0.03, $k$=4) ($p$=0.102) (Figure S7a). There was substantial heterogeneity at $I^2$=69.3%.

Figure S7a: Forest plot for depression

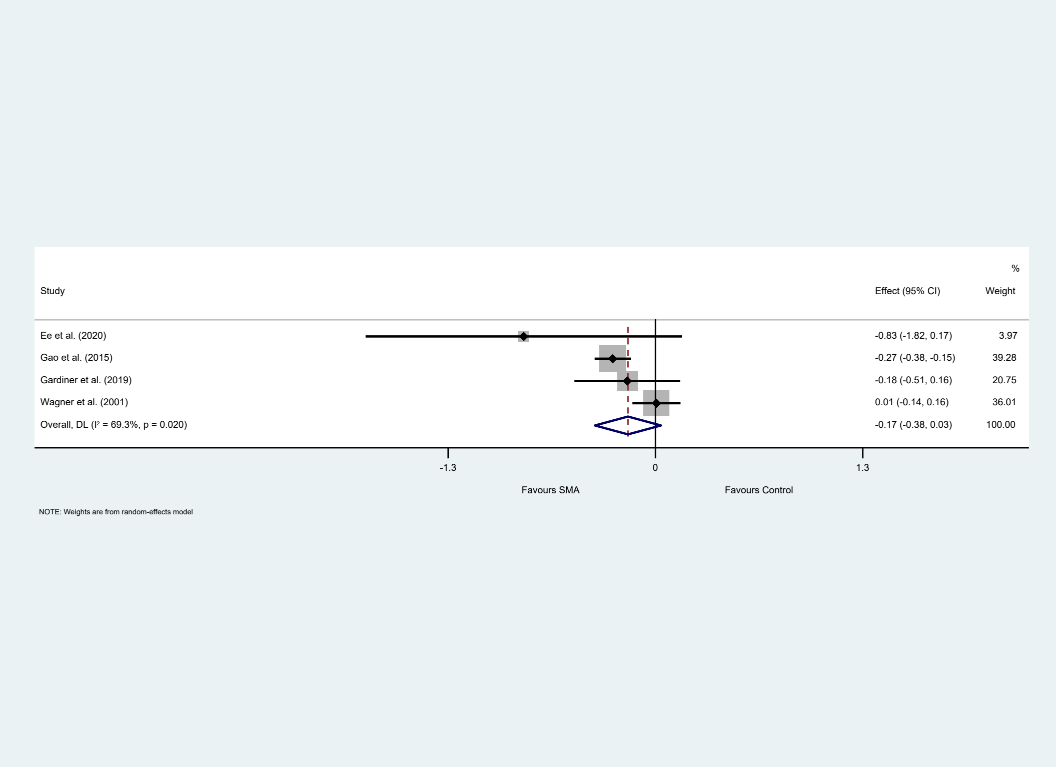


Liu et al. (2012) and Taveira et al. (2011) did not find any differences in depression scores between the SMA and groups [32,43].

### S8 – Additional behavioural outcomes examined with forest plots

*Medication adherence*

Four trials reported on medication adherence [40,46,50,52] but none reported significant differences between the SMA and usual care groups at follow-up.

*Physical activity*

Five SMA trials for metabolic LTCs [23,31,34,40,50] reported physical activity/exercise outcomes, with only Berry et al. (2016) reporting significant between group differences. Berry et al. (2016) measured patient participation (*n*= 80, sample majority non-Hispanic Black females) in five forms of physical activity during the past week: stretching and strengthening exercises, walking, swimming or aquatic exercises, bicycling, and other aerobic exercises. There were no significant differences found for any of these exercises, except patients in the SMA group engaged in more stretching and strengthening exercises (*M*=1.3, *SD*=1.2), compared to patients in the control group (*M*=0.4, *SD*=0.8) [23].

### S9 – SMA delivery, dose and components

#### Mode of SMA delivery

In three trials [33,41,52] the intervention was delivered by a single healthcare professional. In the majority of trials, SMAs were delivered by multidisciplinary teams of interventionists; 15 trials included family physicians [19,23,24,30–34,39,44,46,50,51,53], two trials included specialist diabetes nurse [25,43], five trials included nurse practitioners [23,26–29,32,44,45], seven trials included nurses [19,20,39,40,42,46–50,53] and one trial included a non-specific *'prescribing clinician'* [21,22]. Allied health professionals were involved in the intervention delivery; nine trials included pharmacists [19,20,39,41–43,46–49,52,53], pharmacist with diabetes specialism [40] five trials included dietitians [19,20,29,40,43–45], three trials included physical therapists [19,20,40,42], two involved community health workers [37,38,50], social workers[19,20,53] and or meditation teacher/non prescriber trained in mindfulness or yoga [21,22,31]. Other trials included nutritionists [29,42], healthcare assistants [53], diabetes health educators [31,34–36,44–49] a health coach [30] a registration clerk [53], patient representative group leader [32] and 'trained interventionist' [23]. It was not possible to tell what role each member of staff had in the delivery of the SMA, and whether it was the same of staff to deliver the SMAs each time. The individual consultation component of the intervention was conducted by a physician and/or nurse in twelve trials,[19,22–24,26,31,34,37–39,50,51] pharmacists in three trials [41–43] or was unspecified 'members of the team/clinician' in other studies [32,33,52] (see Table S9).

Provider characteristics other than profession or role were rarely reported. One trial of SMAs for patients with hypertension reported that the physician was female and self-employed [51]. Two trials for diabetic patients involving a majority Hispanic/Latino participants reported that the community health worker and or physician were bilingual, English-Spanish speaking [34,38].

Provider training was reported in several trials and included either 'on the job' training [26,52] and or attendance at a workshop led by those experienced in delivering SMAs. These workshops ranged in duration from 3 hours [27], to one day [32], to 3.5 days [50]. Length of workshop not reported in a fourth trial [33]. Some studies reported the that some providers had undergone training in group facilitation [46], chronic disease management [37,38] or goal setting [33]. One trial reported that clinicians had the opportunity to observe an SMA [25] or pilot the SMA before the study started [33]. Several trials involving patients with diabetes reported that the provider was a certified diabetes educator [40,42–44,46].

Four trials reported that the same interventionists attended all visits for a particular group [40,44,46,51]. One trial of diabetes SMAs reported that group assignments were maintained for all intervention sessions to facilitator peer interactions and relationships within groups [33]. The consistency in group composition in terms of patient and interventionists was not reported by most studies.

**SMA frequency and dose**

The size of each group session ranged from four [40] to 25 [32] patients per group, the most common group size reported was 6-9 patients per group (see Table S9). No studies reported how patients were allocated the SMA sessions (only how they were allocated treatment arm), therefore it is assumed that the groups were mixed in terms of gender, age, ethnicity. A trial of SMAs for older patients reported that family member or carers were encouraged to attend the SMAs sessions [19,29], though it is unclear whether these were excluded from the total number of participants reported or not.

The frequency and duration of the intervention ranged from a single SMA [52], to a series of 16 sessions over the study period [44,45]. Of the trials in which more than one SMA was held, most were conducted monthly [19,25,26,29,30,32,34,37,38], some were held weekly [22,43] others were held weekly to begin with then monthly [40,42]. One trial held SMAs bi-weekly [31] and one trial held SMAs tri-weekly [33]. Two trials held SMAs bi-monthly [46,51], one trial held SMAs tri-monthly [23]. Others trials reported greater variability in frequency; every 3-6 weeks [41], intensive sessions followed by tri monthly sessions [50] or SMAs every 3-6 months [39].

It is difficult to summarise the overall session duration (exposure) as some studies include the arrival and introductions as part of the session and others do not, similarly some studies include the 1:1 consultations as part of the group sessions and others do not. The duration of the sessions reported were typically between 90 [29,34,40,52] and 120 minutes [19,26,30,31,42,43,46,50] however they varied considerably between trials (See Table 3). The shortest duration reported was 30 minutes [41] though this excludes the time taken for the individual consultation at the end of the session. The longest SMA reported was 'half a day' [31] though these occurred less frequently (every 3-6 months). Four studies did not report the length of the SMA sessions [23–25,33,44,45,53].

***SMA intervention components***

Details of intervention components and mode of delivery reported by studies varied. See Table eS9. SMAs were described as 'group medical visits' [23,37,38], or 'group visits (cooperative healthcare clinic)' [19–22,24–28,32,34–36,39,41,42,44,45,50,51] in thirteen trials, 'SMAs' in five trials [29–31,40,53] 'group clinics' [33,46] in two trials, 'chronic care clinic' [39] and 'group medical appointments' [43] and 'group consultations' [52] in one trial, respectively.

A key feature of all SMA interventions was the opportunity for each patient to have an individual 1:1 consultation with a clinician for one or a combination of the following exercises: physical assessment [26,30], medication/prescription review/ adjustments [22,23,25,26,33,34,37,38,42–44,46,50], goal setting [30,46], counselling [39,42], immunisations [24,26], discussion of issues [19,20,31,50,52] review health indicators or goals [29,32,33,40]. These 1:1 consultations occurred at every group session or at least one of the group sessions in a series of sessions (e.g. session 2 [30]).

In most trials (*n*=13) these individual assessments were conducted / offered to all participants during the group session [22,26,29,31,33,34,37–39,41–43,46,51]. In two trials this component was optional, or offered 'as needed' at the very end of the group session [32,50] or immediately after the group session had ended [19,20,24,25,52]. The option to have the individual assessments conducted privately away from the rest of the group members was specified in 3 trials [25,26,52]. Rather than having a 1:1 consultation immediately after the group session, in one trial telephone contact was made with selected patients on an as needed basis to follow-up lab values, self-care monitoring or medication changes [41].

Another feature common to fifteen trials was a facilitated group discussion, or group question and answer session [19,20,23,24,26,29,30,32–34,37–39,50–52]. 'Group education' component was described in thirteen trials [19,23,25,31–33,37,38,42,43,46,50] of which five involved presentations/ lectures from the healthcare professionals [19,24,41–43]. One study of SMAs for diabetes and cardiovascular disease risk included demonstration and coaching of self-care skills [41].

Individual goal setting or action planning was described in 12 trials: lifestyle goals [29,31,41,43,46–49], including dietary goals [40], weight goals [44] health related goals [30], self-management action plans [32–36,41] or therapeutic action plans [42].

Health data collection, including supervised self-assessments, was a component of six SMA interventions. In trials of diabetes SMAs, this included blood glucose self-assessment [46], the collection of 'lab and vitals' [37,38], or health self-assessments [30]. Patient-measured bone fracture risk calculations (FRAX) were collected in the SMA intervention for osteoporosis [52], vital signs, mood state and pain levels were taken by patients at the start of an SMA for chronic pain and depression [22].

Written information provision in the form of handouts was provided in four trials outlining information about the condition [31,52] or nutritional advice [44]. In two trials patients were provided with meditation logbooks [31], or personal risk report cards [41].

Opportunity to socialise was specifically mentioned in eleven trials either by way of a warm-up at the start of the SMA [19,25,26,29,32,34,46,50] or a during a meal towards the end [22,37,38]

Table S9 – Description of SMA intervention composition, duration and components

| Trial | Characteristics of providers (roles within SMA) | Patients per group (n) | Session frequency (n) and each session duration (mins) | SMA components | | | | | | | | |
|---|---|---|---|---|---|---|---|---|---|---|---|---|
| | | | | 1:1 consultation during group | 1:1 consultation after group | Socialisation | Facilitated group discussion/Q&A | Presentation/ education | Written information | Individual goal setting/ action planning | Tools for self-monitoring | self-health assessments/ data collection |
| Scott (2004)[19] | General practitioner/family physician, general nurse, pharmacist, occupational therapist, physiotherapist and dietitian. Receptionist handled scheduling | 8-12 (+caregivers and spouses were invited to attend) 7.7 attended each CHCC group (mean) | Monthly for 24 months, 150 mins | ✓ | | ✓ | ✓ | ✓' | | ✓ | ✓ | ✓ |
| ***Gardiner (2019)***[22] | Prescribing clinician (facilitator) non-prescriber trained in Mindfulness Based Stress Reduction (MBSR) or yoga (co-facilitator) | NR | 9 'in-person' sessions, (weekly over 9 weeks), session duration: 150 mins, Electronic resource access only (over 12 weeks), 10th 'in person' session in week 21, session duration: 150 mins | ✓ | | ✓ | ✓ | | | | | ✓ |
| **Berry (2016)**[23] | Adult health nurse practitioner, a physician, a postdoctoral | NR | 5 sessions, | ✓ | | | | ✓p | | | | |

| | | | | | | | | | | |
|---|---|---|---|---|---|---|---|---|---|---|
| | fellow (clinicians) trained interventionist (facilitator) | | tri- monthly, over 15 months, duration: NR | | | | | | | |
| Clancy (2003)[25] | Hospital physician and specialist nurse (roles NS) | 19-20 | Monthly for 6 months duration: NR | | ✓ | ✓ | ✓ | ✓p | | |
| Clancy (2007)[26] | Primary care internal medicine physicians, registered nurses (roles NS) | 14-17 | Monthly for 1 year, 2 hours | ✓private | | ✓ | ✓ | | | |
| Cole (2013)[29] | Nutrition technician (screener) a dietitian or nutrition technician (session recorder) a certified diabetes educator registered dietitian (provider) and a behavioural specialist, registered nurse or registered dietitian trained in group dynamics (facilitator) | 6-8 (+family members, friends, and other sources of social support) | Monthly over 3 months, 90 mins | ✓ | | ✓ | ✓ | | ✓ | |
| ***Drake (2018)***[30] | Family medicine physician, health coach | 7 or 12? | 8 sessions, approx. monthly over 7 months, 120 mins | ✓ | | | ✓ | | ✓ | ✓ |

| | | | | | | | | | | | |
|---|---|---|---|---|---|---|---|---|---|---|---|
| ***Ee (2020)***[31] | GP, accredited diabetes educator, Meditation teacher | 6-12 | 6 sessions, Bi-weekly, over 3 months, 120 mins | ✓ | | | | ✓ props | ✓ | ✓ | |
| Liu (2013)[32] | General practitioner/family physician, preventive doctor and general nurse practitioner (group facilitators- they alternated leading the 12 group self-management education sessions based on their areas of expertise). Patient rep leader to help coordinate implementation. | 20-25 | Monthly for 12 months Session duration: 150 mins | | ✓selected patients only | ✓ | ✓ | ✓ | | ✓ | |
| Naik (2011)[33] | Primary care physicians (role NS) | 5-7 | Four visits, every 3 weeks 70mins | ✓ | | | ✓ | ✓unclear | | ✓ | |
| Schillinger (2008) [34] | language-concordant physician and health educator (roles NS) | 6-10 | Monthly for 9 months Session duration: 90mins | ✓ | | ✓ | ✓ | | | ✓ | |
| ***Vaughan (2017)***[37] | Physician (clinician) Community Health workers integrated into leadership team (facilitator) CHW selected had '*personality characteristics of flexibility* | 25 participants at session (split into 3 smaller groups during session) | 6 sessions, Monthly, Over 5 months, Session duration: 180mins | ✓ | | ✓ | ✓ | ✓'large group education' | | | ✓ |

| | | | | | | | | | | | | |
|---|---|---|---|---|---|---|---|---|---|---|---|---|
| | *compassion, and determination'* | | | | | | | | | | | |
| ***Vaughan (2020)***[38] | Bilingual physician (clinician) community health worker (group lead) CHW self-identified as Latino, fluent Spanish, CHW certification | NR | Monthly for 6 months Session duration: 3 hours | ✓ | | ✓ | ✓ | ✓'large group education' | | | | ✓ |
| *Wagner (2001)*[39] | Primary care practitioner, nurse and clinical pharmacist (roles NS) | 6-10 | Every 3-6 months, session duration: half-day | ✓ | | | ✓ | | | | | |
| Cohen (2011)[40] | Clinician pharmacist-nationally certified diabetes educator, dietitian, nurse and physical therapist (educator) | 4-6 | 4 once-weekly 2-hour sessions and 5 monthly sessions of 90mins | ✓unclear | | | ✓ | | ✓ | ✓ | | |
| ***Taveira (2014)***[41] | Clinical pharmacist (role NS) | 6-8 | Once every 2-6 weeks until CV risk goal attainment reached. Patients that attained goals followed every 3 months (over 12 months) Session duration; 30 mins | | ✓ | | ✓ | ✓ | ✓ | ✓ | ✓ | |
| ***Wu (2018)***[42] | Clinical pharmacist, nutritionist, nurse or physical | 4-6 | 8 sessions, (Once weekly for over 4 weeks followed by 4 booster | ✓ | | | | ✓ | | ✓ | | |

| | | | | | | | | | | | | |
|---|---|---|---|---|---|---|---|---|---|---|---|---|
| | therapist (roles NS) | | sessions held once every 3 months for a total of 13 months.) Session duration: 120 mins | | | | | | | | | |
| Taveria (2011)[43] | Specialist nurse, pharmacist and dietitian (roles NS) | 4-6 | Four once weekly, session duration: 120 mins | ✓ | | | ✓? | ✓L | | | ✓ | |
| ***Yancy (2020)***[44] | Physicians (2 general internists, 4 endocrinologists), Registered Dietitian, Registered Nurse, and/or Certified Diabetes Educator. (Roles NS) | 8-15 (aim) | 16 sessions, Bi-weekly over 16 weeks, every 8 weeks thereafter, Session duration: | ✓ | | | | ✓ | ✓ | ✓ | | |
| Edelman (2010)[46] | General internist, a pharmacist and a nurse or certified diabetes educator | 7-9 (6-8) | Every 2 months for seven visits over 12 months, 90-120mins | | ✓ | ✓ | ✓ | ✓E | | ✓ | | ✓ |
| ***Gao (2015)***[50] | GP, nurse, community health worker | 18-20 | 6 intensive sessions (over 3 months) followed by continuous sessions (over 6 months), 120 mins | | ✓selected patients | ✓ | ✓ | ✓ | | | | |

| | | | | | | | | |
|---|---|---|---|---|---|---|---|---|
| **Simon and Sawicki (2015)**[51] | Two female physicians. Both self-employed, work in solo practices, without institutional backing. (Roles NS) | 12 invited. 5-12 attendees (average 9.8) | 6 sessions, bi monthly over 12 months, session duration: 75 mins | √ | | √ | | |
| ***Baqir (2016)***[52] | Pharmacist external to practice (clinician) | 13-25 | One 90 mins | | √selected patients | √ | √ | √ |

CHCC- Cooperative Health Care Clinic, P= presentation, L=lecture, E = education
